# A Bivalent Omicron-containing Booster Vaccine Against Covid-19

**DOI:** 10.1101/2022.06.24.22276703

**Authors:** Spyros Chalkias, Charles Harper, Keith Vrbicky, Stephen R. Walsh, Brandon Essink, Adam Brosz, Nichole McGhee, Joanne E. Tomassini, Xing Chen, Ying Chang, Andrea Sutherland, David C. Montefiori, Bethany Girard, Darin K. Edwards, Jing Feng, Honghong Zhou, Lindsey R. Baden, Jacqueline M. Miller, Rituparna Das

**Author notes:** **Corresponding Author:** Spyros Chalkias, MD, Moderna, Inc., Cambridge, MA, USA 02141-2025.

## Abstract

**Background:** Updated vaccination strategies against acute respiratory syndrome coronavirus 2 (SARS-CoV-2) variants of concern are needed. Interim results of the safety and immunogenicity of the bivalent omicron-containing mRNA-1273.214 booster candidate are presented.

**Methods:** In this ongoing, phase 2/3 trial, the 50-μg bivalent vaccine mRNA-1273.214 (25-μg each ancestral Wuhan-Hu-1 and omicron B.1.1.529 spike SARS-CoV-2 mRNAs) was compared to the authorized 50-μg mRNA-1273 booster in adults who previously received 2-dose primary series of 100-μg mRNA-1273 and a first booster dose of 50-μg mRNA-1273 at least 3 months prior. Primary objectives were safety and reactogenicity, and immunogenicity of 50-μg mRNA-1273.214 compared with 50-μg mRNA-1273. Immunogenicity data 28 days after the booster dose are presented.

**Results:** Four hundred thirty-seven and 377 participants received 50-μg of mRNA-1273.214, or mRNA-1273, respectively. Median time between first and second booster doses of mRNA-1273.214 and mRNA-1273 were similar (136 and 134 days, respectively). In participants with no prior SARS-CoV-2 infection, observed omicron neutralizing antibody geometric mean titers (GMTs [95% confidence interval]) after the mRNA-1273.214 and mRNA-1273 booster doses, were 2372.4 (2070.6−2718.2) and 1473.5 (1270.8−1708.4) respectively and the model-based GMT ratio (97.5% confidence interval) was 1.75 (1.49−2.04). All pre-specified non-inferiority (ancestral SARS-CoV-2 with D614G mutation [D614G] GMT ratio; ancestral SARS-CoV-2 [D614G] and omicron seroresponse rates difference) and superiority primary objectives (omicron GMT ratio) for mRNA-1273.214 compared to mRNA-1273 were met. Additionally, mRNA-1273.214 50-μg induced a potent neutralizing antibody response against omicron subvariants BA.4/BA.5 and higher binding antibody responses against alpha, beta, gamma, delta and omicron variants. Safety and reactogenicity profiles were similar and well-tolerated for both vaccines groups.

**Conclusion:** The bivalent vaccine mRNA-1273.214 50-μg was well-tolerated and elicited a superior neutralizing antibody response against omicron, compared to mRNA-1273 50-μg, and a non-inferior neutralizing antibody response against the ancestral SARS-CoV-2 (D614G), 28 days after immunization, creating a new tool as we respond to emerging SARS-CoV-2 variants.

## INTRODUCTION

Severe acute respiratory syndrome coronavirus 2 (SARS-CoV-2) vaccines are safe and effective against coronavirus disease 2019 (Covid-19). The mRNA-1273 vaccine encoding the Wuhan-Hu-1 (ancestral) spike protein of SARS-CoV-2 demonstrated an acceptable safety profile and efficacy of 93.2% against Covid-19 after a median of 5.3 months following the 2-dose 100-μg primary series immunization in the coronavirus efficacy (COVE) trial.^1,2^

Soon after the beginning of the SARS-CoV-2 pandemic, several variants with spike protein mutations that can confer immunologic escape or enhanced transmissability have emerged, such as the beta (B.1.351) and delta (B.1.617.2) variants. In 2022, omicron (B.1.1.529) and omicron subvariants (BA.2, BA.2.12.1, BA.4, BA.5), the most antigenically-divergent variants to date, outcompeted other variants in the setting of a background of substantial pre-existing population immunity from vaccination, infection or both.^3-7^ Omicron and omicron subvariants continue to cause significant morbidity and mortality and re-infection with omicron or a subvariant is possible, especially in unvaccinated individuals.^8-10^

Immunization with an mRNA-1273 50-μg booster dose increases neutralizing antibody responses against variants and booster doses enhance vaccine effectiveness against Covid-19.^11-16^ Nonetheless, effectiveness of the currently authorized vaccines against omicron is decreased compared to other variants^17-20^ and consequently, a second booster dose was authorized in the US for immunocompromised individuals and adults >50 years of age in the midst of omicron waves.^21,22^

Therefore, SARS-CoV-2 candidate vaccines targeting omicron that can induce more potent and durable as well as broader immune responses are important to enhance protection. We have previously evaluated a modified, bivalent booster vaccine approach^23^ with a vaccine that contained equal amounts of mRNA that encode the spike protein of the ancestral SARS-CoV-2 and of the beta variant. Immunogenicity results of the beta-containing bivalent vaccine demonstrated increased and more durable neutralizing antibody responses against beta and multiple variants, even against variants (delta, omicron) not contained in the vaccine, compared to mRNA-1273.^23^ Here we present the interim analysis results 28 days after a 50-μg booster dose of an omicron-containing bivalent candidate, mRNA-1273.214, from an ongoing safety and immunogenicity phase 2/3 study.

## METHODS

### Trial Oversight and Participants

This is an open-label, ongoing Phase 2/3 study (NCT04927065) to evaluate the immunogenicity, safety, and reactogenicity of mRNA-1273.214, a bivalent booster vaccine, as compared to the currently-authorized mRNA-1273 vaccine in adult participants. Adults who had previously received a 2-dose primary series of 100-μg mRNA-1273 and a first booster dose (50 μg) in the COVE^1,2^ or under US emergency use authorization (EUA), were sequentially enrolled. The first cohort received a single second booster of 50-μg of mRNA-1273 (part F, cohort 2) and the second cohort received 50-μg of the bivalent mRNA-1273.214 vaccine which contained equal amounts of ancestral SARS-CoV-2 and omicron spike protein mRNAs (part G). The mRNA-1273 group serves as a non-contemporaneous within-study comparator. Adults with a known history of SARS-CoV-2 infection within 3 months from screening were excluded from the study (inclusion/exclusion criteria are provided in the supplement).

The trial is being conducted across 23 US sites, in accordance with the International Council for Harmonisation of Technical Requirements for Registration of Pharmaceuticals for Human Use, Good Clinical Practice guidelines. The central institutional review board approved the protocol and consent forms. All participants provided written informed consent.

### Trial vaccine

The mRNA-1273.214 50-μg vaccine contains equal amounts of mRNAs (25 μg of each mRNA sequence) that encode the prefusion stabilized spike glycoproteins of the ancestral SARS-CoV-2 (Wuhan-Hu-1) and the omicron variant (B.1.1.529 [BA.1]). The mRNA-1273 50-μg vaccine (Moderna Covid-19 Vaccine) contains only the mRNA sequence encoding the spike glycoprotein of the ancestral SARS-CoV-2. In both vaccines, mRNAs are encapsulated in lipid nanoparticles (LNPs). The booster doses of mRNA-1273.214 and mRNA-1273 were administered intramuscular at doses of 50-μg of mRNA in a 0.5 mL volume.

### Safety Assessment

The primary safety objective was to evaluate the safety and reactogenicity of 50-μg mRNA-1273.214 and of 50-μg mRNA-1273 when administered as a second booster dose (Table S1 and supplementary methods). Reactogenicity included solicited local and systemic adverse reactions (ARs) that occurred ≤7 days after the booster dose as recorded daily by participants. Unsolicited adverse events (AEs) were recorded by study sites for 28 days post-booster administration. Serious adverse events (SAEs), AEs leading to discontinuation from study vaccine and/or participation, medically-attended AEs (MAAEs) and AEs of special interest (AESIs) are to be recorded by the study sites from day 1 through the entire study period (∼12 months).

### Immunogenicity assessment

The pre-specified primary immunogenicity objectives were to demonstrate 1) non-inferior neutralizing antibody responses (based on geometric mean titer [GMT] ratio and seroresponse rate [SRR] difference) or superior neutralizing antibody responses (GMT ratio) against omicron, and 2) non-inferior neutralizing antibody responses (based on GMT ratio) against the ancestral SARS-CoV-2 with the D614G mutation (ancestral SARS-CoV-2 [D614G]), 28 days after the mRNA 1273.214 50-μg second booster dose (day 29) compared with the mRNA-1273 50-μg second booster dose (Table S1 and statistical methods). The pre-specified key secondary objective was to demonstrate non-inferiority (based on SRR difference) against the ancestral SARS-CoV-2 (D614G) 28 days after the mRNA-1273.214 50-μg second booster dose compared with the mRNA-1273 50-μg second booster dose.

Neutralizing antibody geometric mean titers at inhibitory dilutions 50% (ID50) were assessed in validated SARS-CoV-2 spike-pseudotyped lentivirus neutralization assays. Titers were generated against pseudoviruses containing the SARS-CoV-2 full-length spike proteins for the ancestral SARS-CoV-2 (D614G) or the omicron (B.1.1.529 [BA.1]) variant and against a pseudovirus containing full-length spike protein for omicron subvariants BA.4 and BA.5 (referred to as BA.4/BA.5 because of identical spike sequences between BA.4 and BA.5) were assessed using a research-grade pseudovirus assay. Geometric mean (GM) levels were also assessed in an anti-spike protein binding IgG antibody (bAb) assay (Meso Scale Discovery [MSD]) against the ancestral SARS-CoV-2 (D614G), gamma (P.1), alpha (B.1.1.7), beta (B.1.351), delta [B.1.617.2; AY.4], and omicron (B.1.1.529; BA.1) variants (PPD, part of Thermo Fisher Scientific Vaccines Laboratory Services, Richmond, Virginia). Immunogenicity assays are further described in the supplement.

An exploratory objective was to assess incidences of symptomatic and asymptomatic SARS-CoV-2 infection in both groups (Table S1 and Supplementary Methods). Symptomatic infection was evaluated using the primary case definition in the COVE study^1,2^ as well as a secondary case definition based on the Centers for Disease Control and Prevention (CDC) criteria.^24^ Asymptomatic SARS-CoV-2 infection was defined as a positive reverse-transcriptase polymerase chain reaction (RT-PCR) test or a positive serologic test for anti-nucleocapsid antibody after a negative test at the time of enrollment, in the absence of symptoms.

### Statistical analysis

Safety was evaluated in the safety set consisting of all participants who received the 50-μg mRNA-1273.214 or 50-μg mRNA-1273 second booster doses. Solicited adverse reactions were assessed in the solicited safety set (Supplementary Methods). The numbers and percentages of participants with any solicited local and systemic adverse reactions occurring within 7 days post-boost are provided. Unsolicited AEs, SAEs, severe AEs, MAAEs, AESIs and AEs leading to study discontinuation are also summarized.

The primary immunogenicity objectives were assessed in the per-protocol set for immunogenicity–SARS-CoV-2-negative set, also referred to as primary analysis set, consisting of all participants who received the booster dose, had antibody data available at the pre-booster and day 29 visits with no major protocol deviations and had no evidence of SARS-CoV-2 infection pre-booster.

For the pre-specified primary objectives, a hierarchical testing was pre-specified. Superiority of the antibody response of the second booster dose of 50-μg mRNA-1273.214 compared to the second booster dose of 50-μg mRNA-1273 based on the GMR against omicron was to be tested after the following were demonstrated: 1) non-inferiority of the antibody response of the second booster dose of 50-μg mRNA-1273.214 compared with the second booster dose of 50-μg mRNA-1273 based on the GMR against omicron; 2) non-inferiority of the antibody response of the second dose of 50-μg mRNA-1273.214 compared to the second booster dose of 50-μg mRNA-1273 against omicron based on the difference in SRR; and 3) non-inferiority of the antibody response of the second booster dose of 50-μg mRNA-1273.214 compared to the second booster dose of 50-μg mRNA-1273 based on GMR against the ancestral SARS-CoV-2 (D614G) (Fig. S1 and supplementary methods). The key secondary endpoint of non-inferiority of the antibody response of the second booster dose of 50-μg mRNA-1273.214 compared to the second booster dose of 50-μg of mRNA-1273 against the ancestral SARS-CoV-2 (D614G) based on the difference in SRR, was to be tested if all endpoints for the primary objective were met.

The immunogenicity objectives were to be tested 28 days after the booster dose (day 29) and 90 days after the booster dose (day 91) with a two-sided alpha of 0.025 respectively allocated at each one of the two time points. Non-inferiority is considered met when the lower bound of the 97.5% confidence interval (CI) of GMR is ≥0.67 and the SRR-difference is >-10%. Superiority is considered met when the lower bound of the 97.5% CI of GMR is >1. Superiority of the mRNA-1273.214 antibody response against omicron, compared to mRNA-1273, is considered demonstrated if superiority based on GMR is met at days 29 or 91. The day 29 interim analysis results are presented here.

The GMTs (95% CI) for mRNA-1273.214 and mRNA-1273 booster doses are calculated by using t-distribution of log-transformed antibody titers. An analysis of covariance (ANCOVA) model was performed to assess the difference in antibody responses between mRNA-1273.214 and mRNA-1273 groups, with antibody titers post-booster as a dependent variable, and a group variable (mRNA-1273.214 and mRNA-1273) as the fixed effect, adjusting for age groups (<65, ≥65 years) and pre-booster antibody titers. The GMTs (95% CI) estimated by the geometric least square mean (GLSM) from the model for each group and the GMR (mRNA-1273.214 compared with mRNA-1273) estimated by the ratio of GLSM from the model (97.5% CIs) are provided. The 97.5% CI for GMR was used to assess the differences in antibody responses between groups. Seroresponse at a participant level is defined as a change from <lower limit of quantification (LLOQ) to ≥4 × LLOQ, or at least a 4-fold rise if baseline is ≥LLOQ (supplementary methods). The number and percentage of participants who seroresponded is summarized with 95% CI calculated using the Clopper-Pearson method. The differences of SRR between mRNA-1273.214 and mRNA-1273 groups are calculated with 97.5% CI based on stratified Miettinen-Nurminen method adjusting for age groups.

An analysis of the primary immunogenicity endpoints was also performed in the per-protocol set for immunogenicity (participants with and without evidence of prior SARS-CoV-2 infection pre-booster), using an ANCOVA model, with antibody titers at day 29 post-booster as the dependent variable and the vaccine group variable as the fixed effect, adjusting for age groups (<65, ≥65 years), pre-booster SARS-CoV-2 infection status, and pre-booster titers. The SRR difference between the mRNA-1273.214 and mRNA-1273 groups was calculated with 97.5% CI based on stratified Miettinen-Nurminen method adjusted for the pre-booster SARS-CoV-2 infection status and age group. A pre-planned subgroup analysis of participants with prior evidence of SARS-CoV-2-infection pre-booster was performed using an ANCOVA model to assess neutralizing antibody differences between the mRNA-1273.214 and mRNA-1273 groups based on GMRs with 95% CIs. Lastly, a sensitivity analysis was performed excluding the participants with evidence of SARS-CoV-2-infection after the booster dose.

Observed binding antibody GMTs and 95% CIs against variants were provided. The ANCOVA model, adjusting for age group and pre-booster titers, was used to assess binding antibody level differences between the mRNA-1273.214 and mRNA-1273 groups based on GMRs with 95% CIs.

The number and percentage of participants who had asymptomatic or symptomatic SARS-CoV-2 infection and Covid-events are summarized. All analyses were conducted using SAS Version 9.4 or higher.

## RESULTS

### Trial population

Between February 18^th^ to March 8^th^, 2022 (part F, cohort 2, recipients of a second booster dose of mRNA-1273) and March 8^th^ to March 23^rd^, 2022 (part G, recipients of a second booster dose of mRNA-1273.214), a total of 819 participants were enrolled who had previously received the primary series of 100-μg mRNA-1273 and a first booster dose of 50-μg mRNA-1273, at least 3 months prior to enrollment (Fig. 1). Of these, 197 (44.8%) and 243 (55.2%) of the COVE and US EUA participants respectively were assigned to receive a second booster dose of 50-μg mRNA-1273.214 (n=440) and 264 (69.7%) and 115 (30.3%) to receive 50-μg mRNA-1273 (n=379). A total of 437 (53.6%) in the 50-μg mRNA-1273.214 and 377 (46.3%) participants in the 50-μg mRNA-1273 groups, respectively received second boosters. Two (0.5%) participants withdrew consent and discontinued the study after receiving the mRNA-1273.214 booster.

**Figure 1:**
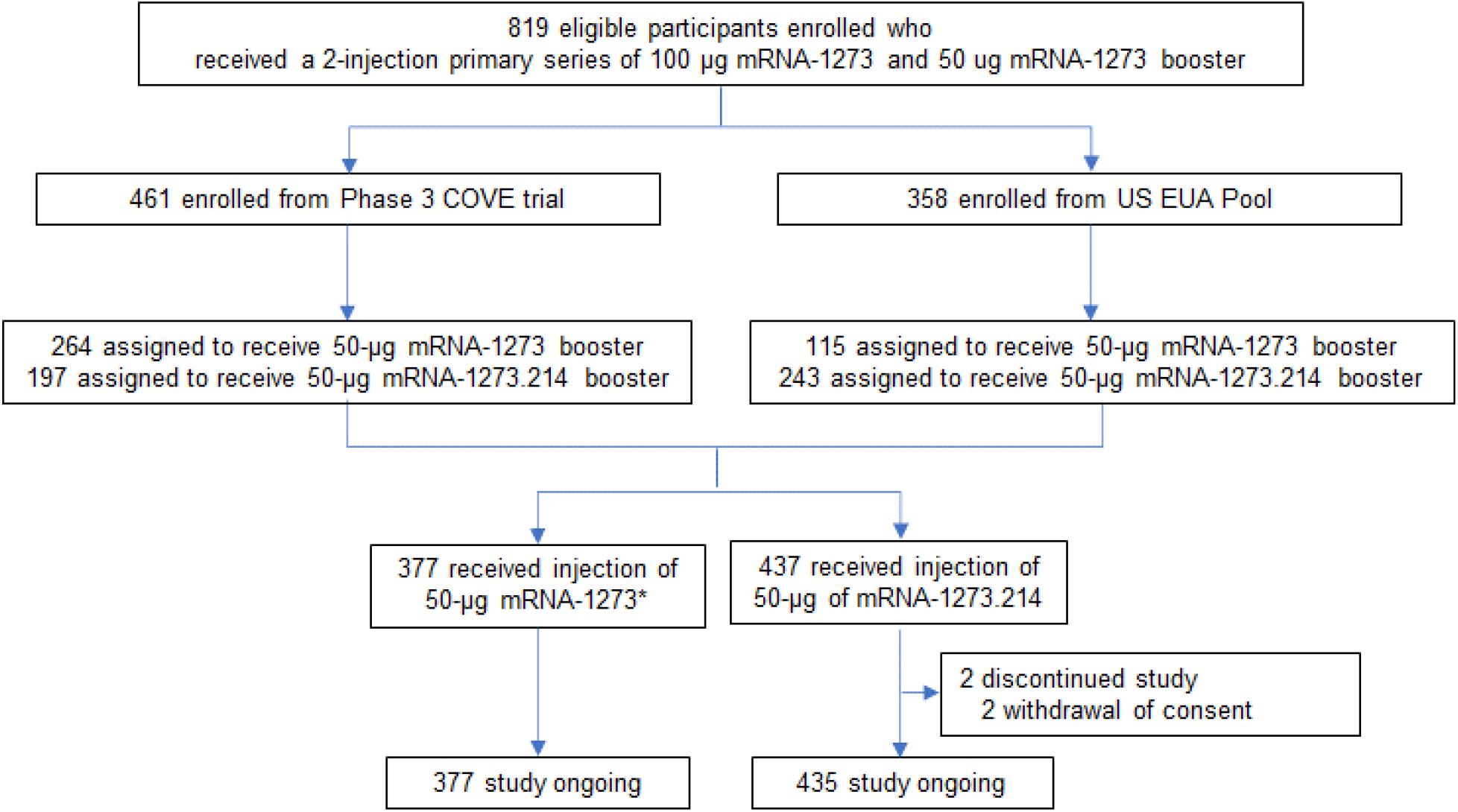
Trial Profile. Emergency Use Authorization, EUA; Coronavirus Efficacy, COVE. Eligible participants who received a prior 2-dose primary series of 100 μg mRNA-1273 and a 50-μg mRNA-1273 booster dose either in the COVE trial or under the US EUA were enrolled to receive a second booster dose of 50-μg mRNA-1273 or mRNA-1273.214. *379 participants were enrolled and dosed; one participant had previously received the primary series but not a first booster dose and another participant had a major protocol deviation, and both were excluded from all analysis sets. Data cutoff date was April 27, 2022.

Participant demographics and baseline characteristics were similar between the two groups (Table 1). The mean ages of the participants in the 50-μg mRNA-1273.214 and 50-μg mRNA-1273 groups, respectively, were 57.3 and 57.5 years, and 59% and 51% were female. Most participants were White (87% and 85%) and 11% and 10% were of Hispanic or Latinx ethnicity in the mRNA-1273.214 and mRNA-1273 groups, respectively. The percentages of participants with evidence of prior SARS-CoV-2 infection (day of the second booster dose) were 22% in the mRNA-1273.214 and 27% in the mRNA-1273 groups. The median time [days (IQR)] between second doses of mRNA-1273 in the primary series and the first booster of mRNA-1273 were similar (245 [224-275] and 242 [225-260]) in the mRNA-1273.214 and mRNA-1273 groups respectively. Median time (days [IQR]) between the first booster dose of mRNA-1273 and the second booster doses were similar (136 [118-150] and 134 [118-150]).

**Table 1.** Demographics and Study Participant Characteristics.

| Characteristics n (%)* | mRNA-1273.214<br>(50 µg), N=437 | mRNA-1273<br>(50 µg), N=377 |
| --- | --- | --- |
| Age at Screening (yr) |  |  |
| Mean (range) | 57.3 (20, 88) | 57.5 (20, 96) |
| Age subgroup |  |  |
| ≥18 and <65 years | 263 (60.2) | 227 (60.2) |
| ≥65 years | 174 (39.8) | 150 (39.8) |
| Gender |  |  |
| Male | 179 (41.0) | 186 (49.3) |
| Female | 258 (59.0) | 191 (50.7) |
| Ethnicity |  |  |
| Hispanic or Latino | 46 (10.5) | 37 (9.8) |
| Not Hispanic or Latino | 390 (89.2) | 340 (90.2) |
| Not reported or unknown | 1 (0.2) | 0 |
| Race |  |  |
| White | 381 (87.2) | 322 (85.4) |
| Black or African American | 31 (7.1) | 29 (7.7) |
| Asian | 14 (3.2) | 16 (4.2) |
| American Indian or Alaska Native | 0 | 1 (0.3) |
| Native Hawaiian or Other Pacific Islander | 0 | 1 (0.3) |
| Multiracial | 7 (1.6) | 2 (0.5) |
| Other | 3 (0.7) | 2 (0.5) |
| Not reported or unknown | 1 (0.2) | 4 (1.1) |
| Time between second injection of mRNA-1273 in the primary series and the first booster of mRNA-1273 (days) |  |  |
| n | 435 | 374 |
| Median | 245.0 | 242.0 |
| IQR | 51.0 | 35 |
| Time between first booster injection of mRNA-1273 and the second booster (days) |  |  |
| n | 435 | 374 |
| Median | 136.0 | 134.0 |
| IQR | 32 | 32 |
| Pre-booster RT-PCR SARS-CoV-2 |  |  |
| Negative | 434 (99.3) | 367 (97.3) |
| Positive | 2 (0.5) | 2 (0.5) |
| Missing | 1 (0.2) | 8 (2.1) |
| Pre-booster antibody to SARS-CoV-2 nucleocapsid† |  |  |
| Negative | 341 (78.0) | 276 (73.2) |
| Positive | 95 (21.7) | 100 (26.5) |
| Missing | 1 (0.2) | 1 (0.3) |
| Pre-booster SARS-CoV-2 status‡ |  |  |
| Negative | 340 (77.8) | 267 (70.8) |
| Positive | 96 (22.0) | 101 (26.8) |
| Positive by both RT-PCR and SARS-CoV-2 nucleocapsid† | 1 (0.2) | 1 (0.3) |
| Positive by RT-PCR only | 1 (0.2) | 1 (0.3) |
| Positive by SARS-CoV-2 nucleocapsid only† | 94 (21.5) | 99 (26.3) |
| Missing | 1 (0.2) | 9 (2.4) |
| IQR=interquartile range; RT-PCR = reverse transcription polymerase chain reaction. *Percentages based on the number of participants in the safety set. †Elecsys assay for binding antibody to SARS-CoV-2 nucleocapsid. ‡Pre-booster SARS-CoV-2 status was positive if there was evidence of prior SARS-CoV-2 infection, defined as positive binding antibody against the SARS-CoV-2 nucleocapsid or positive RT-PCR at Day 1; Negative SARS-CoV-2 status was defined as negative binding antibody against the SARS-CoV-2 nucleocapsid and a negative RT-PCR at Day 1. Data cutoff date is April 27<sup>th</sup>, 2022. |  |  |

### Safety

Median durations of follow-up days (IQR) were 43 (41-45) for the mRNA-1273.214 and 57 (56-62) for the mRNA-1273 boosters; the difference is due to the sequential nature of group assignment. The occurrence of solicited local adverse reactions (ARs) within seven days following second booster doses was 79% in mRNA-1273.214 and 80% in mRNA-1273 (Fig. 2 and Table S3). The most commonly reported solicited local adverse reaction within 7 days after administration of both second boosters was injection site pain for both groups (77%). The incidences of systemic ARs were 70% in the mRNA-1273.214 and 66% in the mRNA-1273 groups and the most frequent ARs were fatigue (55% and 51%), headache (44% and 41%), myalgia (40% and 39%), and arthralgia (31% and 32%) in these groups, respectively. The majority of solicited ARs were mild-to-moderate (grades 1-2) for both groups. The frequency of grade 3 events was 8% in both groups, and the most commonly reported grade 3 ARs were fatigue (both 3%), and myalgia (2% and 4%) for the mRNA-1273.214 and mRNA-1273 groups, respectively. No grade 4 events occurred in either group.

**Figure 2.**
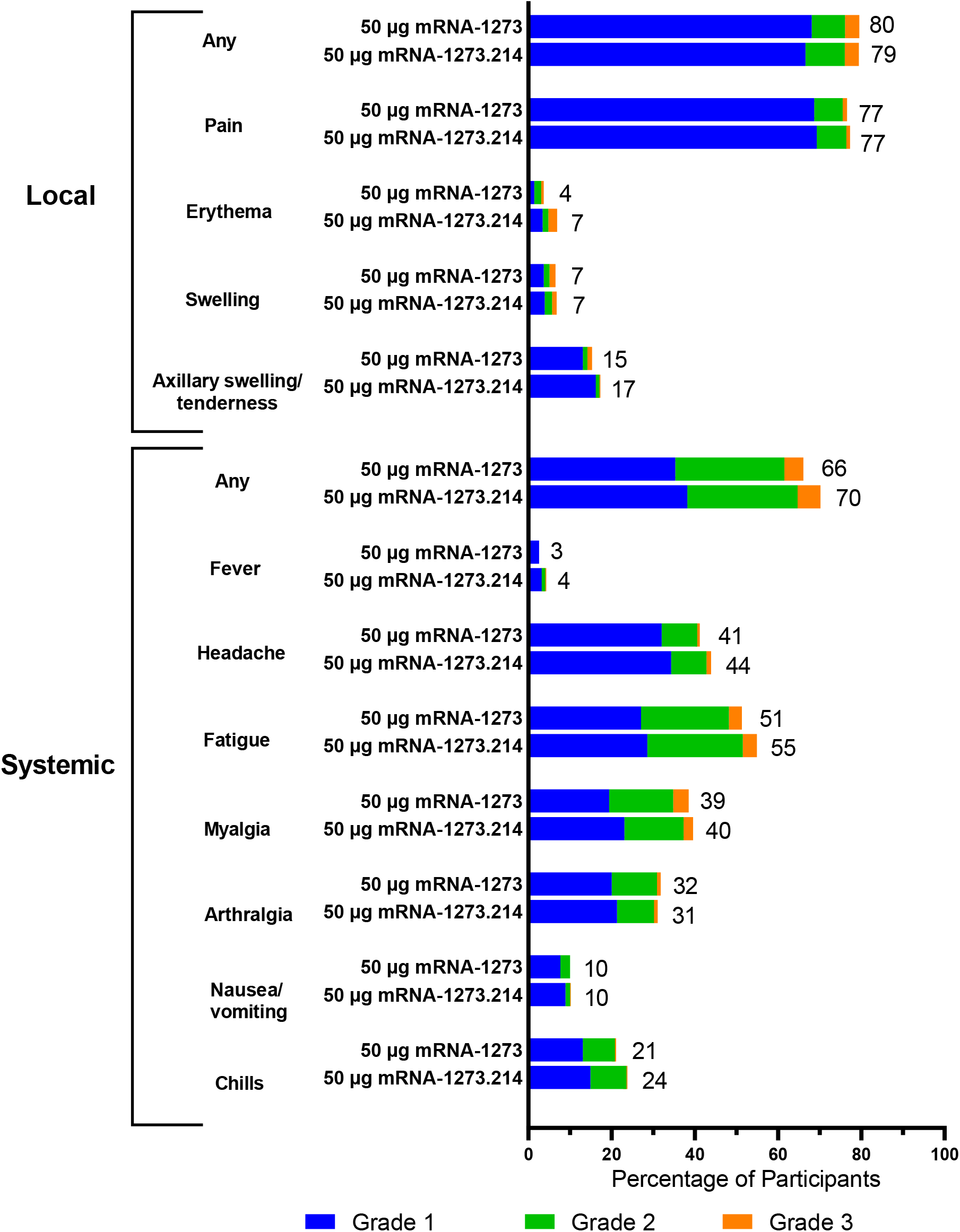
Solicited Local and Systemic Adverse Reactions. Percentages of participants who had solicited local or systemic adverse reactions within 7 days after the booster dose by grade.

Unsolicited adverse events regardless of the relationship to vaccination up to 28 days after the second booster doses were reported in 19% of participants in the mRNA-1273.214 and 21% in the mRNA-1273 groups (Table S4). The overall incidences of AEs considered related to the study vaccination by the investigator was 6% in both booster groups. In the mRNA-1273.214 group, two participants experienced serious adverse events (SAEs, prostate cancer and traumatic fracture) and one participant in the mRNA-1273 group reported an SAE of spinal osteoarthritis within 28 days of the booster dose; none were assessed by site investigators to be related to study vaccination. Medically-attended adverse events (MAAEs) were 10% in the mRNA-1273.214 and 14% in the mRNA-1273 groups up to 28 days after vaccination, and of those, 2 participants (1%) had MAAEs considered related to study vaccination in the mRNA-1273.214 (grade 2, fatigue and grade 1, dermatitis) and mRNA-1273 (hypertension and urticaria, both grade 1) groups. There were no fatal events or AEs leading to study discontinuation. As of the data cutoff date, no deaths and no events of myocarditis or pericarditis occurred, and one additional SAE (grade 3, nephrolithiasis), considered unrelated to study vaccination by the investigator, was reported in the mRNA-1273.214 group.

### Immunogenicity

In the primary analysis set of participants without evidence of prior SARS-CoV-2, the observed neutralizing antibody GMTs (95% CI) against the ancestral SARS-COV-2 (D614G) were 5977.3 (5321.9-6713.3) and 5649.3 (5056.8-6311.2) and the omicron GMTs were 2372.4 (2070.6-2718.2) and 1473.5 (1270.8-1708.4) 28 days after the mRNA-1273.214 and mRNA-1273 doses, respectively (Table 2). Estimated GMTs in the primary analysis set after adjusting for age groups and pre-booster titers were 6422.3 (5990.1−6885.7) and 5286.6 (4887.1−5718.9) against the ancestral virus (D614G) 28 days after the mRNA-1273.214 and mRNA-1273 booster doses, respectively, with a GMR (97.5% CI) of 1.22 (1.08−1.37), meeting the pre-specified criterion for non-inferiority (lower bound of CI ≥0.67). The estimated GMTs against omicron were 2479.9 (2264.5−2715.8) and 1421.2 (1283.0−1574.4) 28 days following the mRNA-1273.214 and mRNA-1273 booster doses, respectively, with a GMR (97.5% CI) of 1.75 (1.49−2.04) which met the pre-specified superiority criterion (lower bound of CI >1).

**Table 2.** Primary Immunogenicity Analysis of Ancestral SARS-CoV-2 (D614G) and Omicron after 50-μg of mRNA-1273.214 and mRNA-1273 Administered as Second Booster Doses.

|  | Ancestral SARS-CoV-2 (D614G) |  | Omicron |  |
| --- | --- | --- | --- | --- |
|  | 50 µg<br>mRNA-1273.214<br>Booster Dose<br>N=334 | 50 ug<br>mRNA-1273<br>Booster Dose<br>N=260 | 50 µg<br>mRNA-1273.214<br>Booster Dose<br>N=334 | 50 ug<br>mRNA-1273<br>Booster Dose<br>N=260 |
| Pre-booster n† | 334 | 260 | 334 | 260 |
| Observed GMT (95% CI)§ | 1266.7<br>(1120.2-1432.5) | 1521.0<br>(1352.8-1710.2) | 298.1<br>(258.8-343.5) | 332.0<br>(282.0-390.9) |
| Day 29, n† | 334 | 260 | 334 | 260 |
| Observed GMT (95% CI)§ | 5977.3<br>(5321.9-6713.3) | 5649.3<br>(5056.8-6311.2) | 2372.4<br>(2070.6-2718.2) | 1473.5<br>(1270.8-1708.4) |
| GMFR (95% CI)§ | 4.7 (4.4-5.1) | 3.7 (3.4-4.0) | 8.0 (7.2-8.8) | 4.4 (4.0-5.0) |
| Estimated GMT (95% CI)¶¶ | 6422.3<br>(5990.1-6885.7) | 5286.6<br>(4887.1-5718.9) | 2479.9<br>(2264.5-2715.8) | 1421.2<br>(1283.0-1574.4) |
| GMR (97.5% CI)¶¶\|\| | 1.22 (1.08, 1.37) |  | 1.75 (1.49-2.04) ¶¶ |  |
| Day 29 SRR, n/N1 %‡<br>(95% CI) | 334/334, 100<br>(98.9-100) | 260/260-100<br>(98.6-100) | 333/333, 100<br>(98.9-100) | 256/258, 99.2<br>(97.2-99.9) |
| Difference (97.5% CI)\|\|†† | 0 |  | 1.5 (-1.1- 4.0) |  |
| ANCOVA=analysis of covariance, CI = Confidence interval, GMT=geometric mean titer; GMFR= geometric mean fold-rise; GMR = Geometric mean ratio, mRNA-1273.214 vs mRNA-1273. ID50, 50% inhibitory dose; LLOQ, lower limit of quantification; LS, least squares; ULOQ, upper limit of quantification. Antibody values assessed by pseudovirus neutralizing antibody assay reported as below the LLOQ (18.5 for ancestral [D614G] and 19.9 for omicron are replaced by 0.5 × LLOQ. Values greater than ULOQ (45,118) for ancestral [D614G] and (15,502.7) for omicron are replaced by the ULOQ if actual values are not available.<br>†Number of participants with non-missing data at the timepoint (baseline or post-baseline).<br>§GMFR of nAb at day 29 post-baseline timepoint over pre-dose day 1 with corresponding 95% CI based on the t-distribution of log-transformed values or difference in the log-transformed values for GMT value and GMT fold-rise, respectively, then back transformed to the original scale.<br>¶ Log-transformed antibody levels are analyzed using an ANCOVA model with the treatment variable as fixed effect, adjusting for age group (<65, ≥65 years) and pre-booster titers. The resulting LS means, difference of LS means, and 95% CI are back transformed to the original scale.<br>¶¶97.5% CI was calculated by stratified Miettinen-Nurminen method adjusted by age group.<br>‡Seroresponse at a participant level defined as a change from <LLOQ to ≥4 × LLOQ, or at least a 4-fold rise if baseline is ≥LLOQ; comparison to pre-vaccination baseline. Percentages were based on the number of participants with non-missing data at baseline and the corresponding time point, 95% CI calculated using the Clopper-Pearson method.<br>††The SRR difference is a calculated common risk difference using inverse-variance stratum weights and the middle point of Miettinen-Nurminen confidence limits of each one of the stratum risk differences. The stratified Miettinen-Nurminen estimate and the CI cannot be calculated when the seroresponse rate in both groups is 100%, absolute difference is reported.<br>¶¶¶ Exceeded non-inferiority criteria and met superiority criteria including lower bound CI >1 and testing sequence. |  |  |  |  |

The seroresponse rates (SRRs) (95% CI) against ancestral SARS-COV-2 (D614G) were 100% (98.9−100% and 98.6−100%) 28 days after the mRNA-1273.214 and mRNA-1273 booster doses, respectively, with an SRR difference of 0 meeting the non-inferiority criterion (lower bound of CI >-10%). The omicron SRRs were 100% (98.9−100%) for mRNA-1273.214 and 99.2% (97.2−99.9%) for mRNA-1273 28 days following the booster doses, and the estimated SRR difference (97.5% CI) was 1.5% (−1.1−4.0) meeting the non-inferiority criterion (lower bound of CI >-10%). Therefore, all primary and key secondary immunogenicity endpoints were met based on the pre-specified testing sequence (Fig. S1). All immunogenicity endpoints were also met based on the analysis which included all participants, with and without evidence of SARS-CoV-2 infection pre-booster (Table S5).

To evaluate whether the antibody responses were consistent between participants with and without prior SARS-CoV-2 infections, a pre-planned subgroup analysis was performed (Fig. 3 and Fig. S2; Tables S6-S7). In those with evidence of prior SARS-CoV-2 infection, observed GMTs against the ancestral SARS-CoV-2 (D614G) were 9509.7 (7345.9-12310.9) and 7003.5 (5592.6-8770.4) and estimated GMTs were 9891.5 (8732.2-11204.8) and 7776.5 (6813.0-8876.3) for the mRNA-1273.214 and mRNA-1273 groups, respectively with a GMR (95% CI) of 1.27 (1.07-1.51). Against omicron, observed GMTs (95% CI) were 7676.2 (5618.2-10488.1) and 3885.6 (2877.8-5246.4) and estimated GMTs were 7669.2 (6470.7-9089.6) and 4041.5 (3375.1-4839.5) for mRNA-1273.214 and mRNA-1273 with a GMR (95% CI) of 1.90 (1.50-2.40). For both groups, the SRRs were 100% for the ancestral SARS-CoV-2 (D614G) and omicron, and the SRR difference was 0.

**Figure 3.**
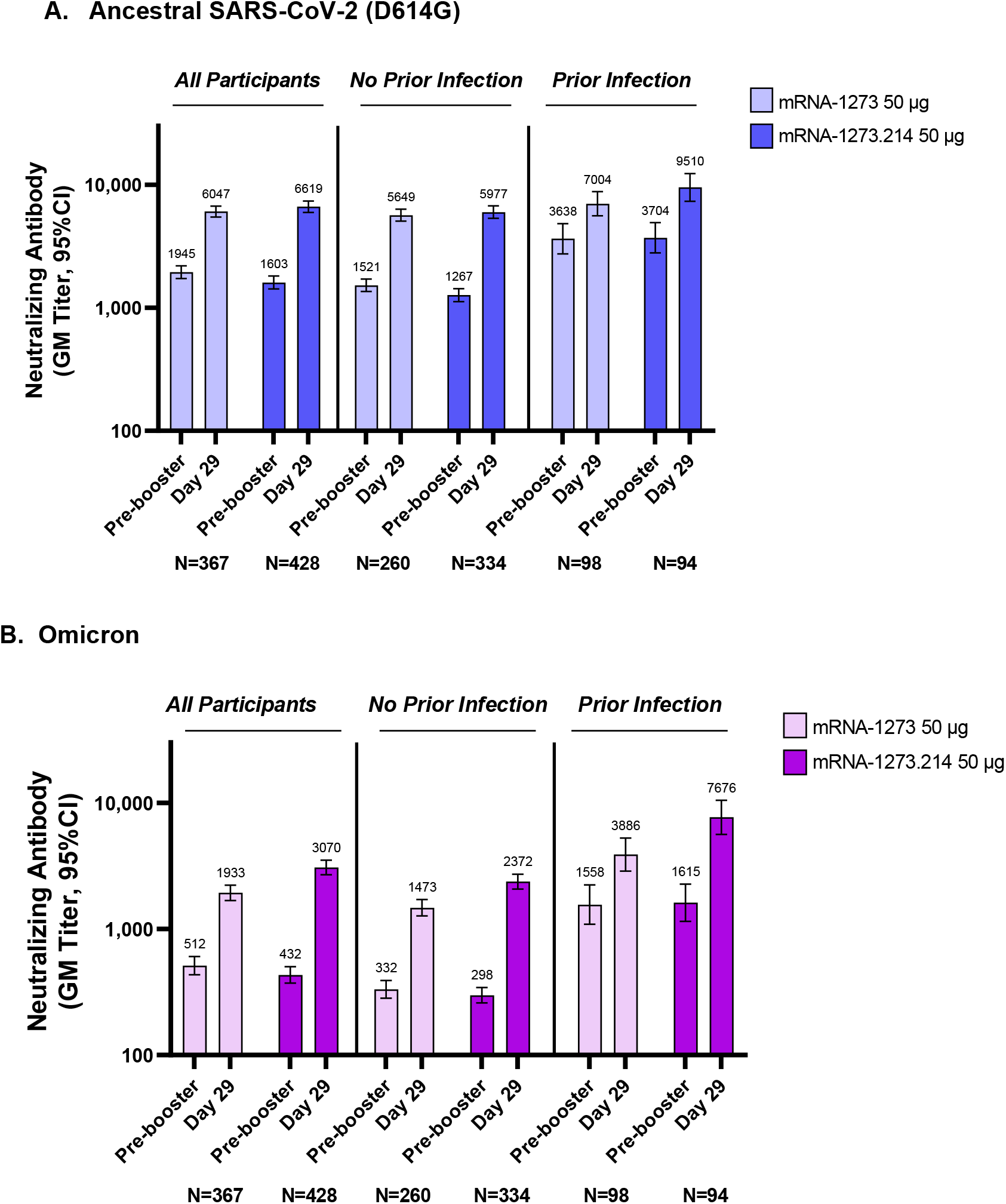
Observed Neutralizing Antibody Titers Against Ancestral SARS-CoV-2 (D614G) and Omicron after 50-μg of mRNA-1273.214 and mRNA-1273 Administered as Second Booster Doses. GM = geometric mean; CI = confidence interval. Pseudovirus neutralizing antibody geometric mean titers are from participants with non-missing data at the timepoint. Nine participants in the mRNA-1273 arm were missing pre-booster SARS-CoV-2 status. Antibody values reported as below the lower limit of quantification (LLOQ: 18.5 for ancestral SARS-CoV-2 (D614G); 19.9 for omicron) were replaced by 0.5 x LLOQ. Values greater than the upper limit of quantification (ULOQ: 45,118 for ancestral SARS-CoV-2 (D614G); 15502.7 for omicron) were replaced by the ULOQ if actual values are not available. 95% CIs were calculated based on the t-distribution of the log-transformed values or the difference in the log-transformed values for GM value and GM fold-rise, respectively, then back transformed to the original scale for presentation. Data for observed nAb GMTs by prior infection status are provided in Table S7.

Given the emergence of omicron subvariants, we also assessed the neutralizing antibody response 28 days after the 50-μg mRNA-1273.214 booster dose against the omicron BA.4 and BA.5 subvariants which have additional spike mutations compared to BA.1. The observed GMTs, 28 days following the booster dose, were 727.4 (632.8-836.1) and the fold-rise in titers, compared to the pre-booster level of 115.6 (98.5-135.6) was 6.3 (5.7-6.9) in participants without evidence of prior SARS-CoV-2 infection pre-booster (Fig. S3 and Table S8). In the subgroup of participants with prior SARS-CoV-2 infection, the GMTs against the omicron subvariants was 2337.4 (1825.5-2992.9) and the fold-rise from the pre-booster level of 719.5 (531.6, 973.9) was 3.2 (2.8-3.8). In all participants regardless of prior SARS-CoV-2 infection, GMTs were 172.7 (147.5-202.3) pre-booster and rose to 940.6 (826.3-1070.6) with a fold-rise of 5.4 (5.0-5.9).

Differences between the two vaccines based on binding antibody titers were also assessed for ancestral SARS-CoV-2 and omicron, as well as other variants of concern. In participants without evidence of prior SARS-CoV-2 infection, the binding antibody GMTs (95% CI) were higher (nominal alpha of 0.05) following mRNA-1273.214 than mRNA-1273 booster doses with GMRs (95% CI) of 1.19 (1.10-1.28) for alpha, 1.15 (1.07-1.25) for beta, 1.19 (1.10-1.28) for gamma and 1.11 (1.03-1.19) for delta (Fig. S4 and Table S9). Similar GMRs between the mRNA-1273.214 and mRNA-1273 vaccine groups were observed in the analysis of all participants regardless of prior SARS-CoV-2 infection (Table S10). The observed binding antibody titers for the ancestral SARS-CoV-2 and omicron are summarized in Table S11.

### Incidence of SARS-CoV-2 infections

In participants with no prior evidence of SARS-SoV-2 infection, starting 14 days after vaccination, SARS-CoV-2 infections, regardless of the presence of symptoms, occurred in 11 (3.2%) and 5 (1.9%) participants in the mRNA-1273.214 and mRNA-1273 groups, respectively. Of those 6 (1.8%) and 4 (1.5%) infections were asymptomatic, and there were 5 (1.5%) and 1 (0.4%) Covid-19 events (symptomatic infections) in the mRNA-1273.214 and mRNA-1273 groups, respectively. There were no emergency room visits or hospitalizations due to Covid-19.

## DISCUSSION

Our results indicate that a 50-μg dose of the bivalent omicron-containing mRNA-1273.214 vaccine had a safety and reactogenicity profile that was similar to the prototype mRNA-1273 (50-μg) booster vaccine when administered as second booster doses. The frequency of the adverse reactions after a second booster dose of 50-μg mRNA-1273.214 was similar to or lower than that of a first booster dose of 50-μg mRNA-1273 and of the second dose of the 100-μg mRNA-1273 primary series as previously reported.^11,15^ Overall, the reactogenicity profile of mRNA-1273.214 is reassuring, with three prior doses administered and with an interval of at least 3 months from the prior dose. We have also previously evaluated the safety and reactogenicity of another bivalent candidate, mRNA-1273.211, which had similar safety and reactogenicity to the mRNA-1273.214 vaccine, with a reassuring safety profile through 6 months post-vaccination.^23^

Neutralizing antibody responses have been used as a surrogate to assess effectiveness of Covid-19 vaccines.^25,26^ The 50-μg mRNA-1273.214 booster vaccine elicited a superior neutralizing antibody response against omicron, compared to 50-μg mRNA-1273, 28 days after the booster dose. The magnitude of the difference in the omicron neutralizing antibody response between the two vaccines (GMR 1.75 [97.5% CI, 1.49−2.04]), exceeds the recommended superiority criteria.^27^ The neutralizing antibody response against the ancestral SARS-CoV-2 (D614G) was also higher with mRNA-1273.214, compared to mRNA-1273, indicating no decrement in the ancestral SARS-CoV-2 (D614G) antibody responses after the booster dose of the bivalent vaccine. Neutralizing antibody responses were consistently higher with mRNA-1273.214, compared to mRNA-1273, in participants with and without evidence of prior SARS-CoV-2 infection. Given the epidemiological data suggestive of decreasing vaccine effectiveness against infection with omicron and that break-through infections can occur in vaccinated individuals, including those with prior SARS-CoV-2 infection,^8-10,17,28^ it is important to be able to boost immune responses in this group. The mechanisms of an increased antibody response with vaccines that contain both the ancestral SARS-CoV-2-spike protein sequence as well as that of an antigenically-divergent variant could include the generation of new memory immune responses but are yet to be elucidated.^29^

The ability to cross-react with multiple variants is highly desirable given the continuous evolution of SARS-CoV-2 and the emergence of escape variants. The results of the binding antibody responses indicate higher responses with the bivalent omicron-containing vaccine, compared to the prototype mRNA-1273, even when the variants are not contained in the vaccine. We had previously reported that a bivalent booster vaccine containing the ancestral SARS-CoV-2 and an antigenically-divergent variant can induce antibodies which cross-neutralize and cross-react with multiple variants and consistently elicit higher antibody responses compared to mRNA-1273.^23^ mRNA-1273.214 elicited potent neutralizing antibodies responses against the omicron BA.4 and BA.5 subvariants and the increase (fold-rise) in neutralizing antibody titers was consistent with the increase in titers against omicron BA.1, regardless of prior SARS-CoV-2 infection. In addition, the longevity of the neutralizing antibody response is important and we previously observed a more durable response against multiple variants, 6 months after immunization with the beta-containing bivalent vaccine, compared to mRNA-1273.^23^ We will continue to monitor the persistence of the antibody responses elicited by mRNA-1273.214 in the ongoing clinical study.

There are limitations to the study, including that it was not randomized. While the enrollment dates of the booster groups were within weeks of each other, representing similar epidemiological environments of circulating variants, we did not ascertain variant sequences. Additionally, the temporal distances between primary vaccination and first and second booster doses were similar between the two groups, despite the sequential study design. The study only assessed humoral immune responses; thus, future work is needed to characterize the potential contribution of cellular responses to protection. We present data from day 28 post-boost given the public health importance of these data; longer term follow-up is underway to characterize the antibody persistence and, although the study was not designed to evaluate vaccine effectiveness, to describe infection rates post-boost.

In conclusion, the omicron-containing bivalent vaccine mRNA-1273.214 had a safety and reactogenicity profile similar to that of the current booster vaccine mRNA-1273 when administered at the 50-μg dose. mRNA-1273.214 elicited a superior neutralizing antibody response against omicron, compared to mRNA-1273, and potent neutralizing antibody responses against the BA.4 and BA.5 omicron subvariants 28 days after immunization. Antibody responses were also higher against the ancestral SARS-CoV-2 (D614G) and multiple additional variants. These results are consistent with the evaluation of our first, beta-containing, bivalent vaccine which induced enhanced and durable antibody responses against multiple SARS-CoV-2 variants^23^ and bivalent vaccines can be a new tool as we respond to emerging variants.

## Supporting information

CONSORT Checklist

## Data Availability

As the trial is ongoing, access to patient-level data and supporting clinical documents with qualified external researchers may be available upon request and subject to review once the trial is complete.

## AUTHOR CONTRIBUTIONS

SC, JF, JMM, RD and HZ contributed to the design of the study and oversight. SC, SRW, AB, CH, and NMcG contributed to data collection. BG and DCM were responsible for immunogenicity assays. SC, JF, HZ, RD, JMM, LRB and JET contributed to data analysis and/or interpretation of the data. SC, JF, LRB and JET contributed to drafting the manuscript. All authors critically reviewed and provided input to manuscript drafts and approved the final version for submission to the journal.

## ACKNOWLEGEMENTS

We thank the participants in the trial and the members of the mRNA-1273 trial team (listed in the Supplementary Appendix) for their dedication and contributions to the trial, the Immune Assay Team at Duke University Medical Center, Durham, NC for PsVNA analyses, and Frank J. Dutko, Ph.D., (Moderna consultant) for writing and editorial support.

