## Supplementary Appendix for "A Bivalent Omicron-containing Booster Vaccine Against Covid-19"

**TABLE OF CONTENTS**


| **Contents** | **Pg.** |
|---|---|
| List of Investigators | 3 |
| Supplementary Methods | 4 |
| Study Design | 4 |
| Part G Study Eligibility Criteria | 4 |
| Immunogenicity Objectives and Endpoints | 6 |
| Statistical Analysis of Immunogenicity | 6 |
| Immunogenicity assays | 8 |
| Incidence of SARS-CoV-2 Infection and Covid-19 | 9 |
| Figure S1. Statistical Testing Sequence | 11 |
| Figure S2. Observed Neutralizing Antibody Titers Against SARS-CoV-2 (D614G) and Omicron after 50-µg of mRNA-1273.214 and mRNA-1273 Administered as Second Booster Doses | 12 |
| Figure S3. Observed Neutralizing Antibody Titers Against Omicron BA.4 and BA.5 Subvariants after 50-µg of mRNA-1273.214 Administered as a Second Booster Dose | 14 |
| Figure S4. Observed Binding Antibody Levels Against Variants After 50-µg of mRNA-1273.214 and mRNA-1273 Administered as Second Booster Doses in All Participants | 15 |
| Table S1. Objectives and Endpoints for Part G Second Booster Doses of 50-µg mRNA-1273.214 in Participants Who Received 100 µg mRNA-1273 Primary Series and a Booster Dose of 50 µg mRNA-1273 | 16 |
| Table S2. Analysis Sets | 18 |
| Table S3. Solicited Local and Systemic Adverse Reactions Within 7 Days Following the Second Booster Injections of mRNA-1273.214 (50 µg) and mRNA-1273 (50 µg) | 19 |
| Table S4. Summary of Unsolicited Adverse Events ≤28 Days Post-booster dose | 21 |
| Table S5. Geometric Mean Neutralizing Antibody Titer Ratio and Seroresponse Difference Against Ancestral SARS-CoV-2 (D614G) and Omicron after 50-µg of mRNA-1273.214 and mRNA-1273 Administered as Second Booster Doses (Supportive Analysis – All Participants Regardless of Evidence of Prior SARS-CoV-2 Infection | 22 |
| Table S6. Geometric Mean Neutralizing Antibody Titer Ratio and Seroresponse Difference Against Ancestral SARS-CoV-2 (D614G) and Omicron after 50-µg of mRNA-1273.214 and mRNA-1273 administered as Second Booster Doses in Participants with Evidence of Prior SARS-CoV-2 Infection | 23 |
| Table S7. Observed Neutralizing Antibody Geometric Mean Titers Against Ancestral SARS-CoV-2 (D614G) and Omicron after 50-µg of mRNA-1273.214 and mRNA-1273 Administered as Second Booster Doses by Prior SARS-CoV-2 Infection at Pre-Booster | 24 |
| Table S8. Observed Neutralizing Antibody Geometric Mean Titers Against Omicron BA.4 and BA.5 Subvariants after 50-µg of mRNA-1273.214 Administered as Second Booster Doses by Prior SARS-CoV-2 Infection at Pre-Booster | 25 |
| Table S9. Binding Antibody Titers and Geometric Mean Ratios Against Ancestral SARS-CoV-2 and Variants after 50-µg of mRNA-1273.214 and mRNA-1273 Administered as Second Booster Doses in Participants with No Evidence of Prior SARS-CoV-2 Infection at Pre-Booster | 26 |
| Table S10. Binding Antibody Levels and Geometric Mean Ratios Against Ancestral SARS-CoV-2 and Variants after 50-µg of mRNA-1273.214 and mRNA-1273 | 27 |

| | |
|---|---|
| Administered as Second Booster Dose, Regardless of Evidence of Prior SARS-CoV-2 Infection | |
| Table S11. Observed Binding Antibody Geometric Mean Levels Against Ancestral SARS-CoV-2 and Omicron After Second 50-µg Boosters of mRNA-1273.214 and mRNA-1273 by Prior SARS-CoV-2 Infection at Pre-Booster | 28 |
| Supplement References | 29 |

### List of Investigators

| Investigator | Institution | Location |
|---|---|---|
| Paul Bradley | Meridian Clinical Research | Savannah, Georgia |
| Adam Brosz | Meridian Clinical Research | Grand Island, Nebraska |
| Laurence Chu | Benchmark Research | Austin, Texas |
| Michael Cotugno | Benchmark Research | Metairie, Louisiana |
| Frank Eder | Meridian Clinical Research, LLC | Binghamton, New York |
| David Ensz | Meridian Clinical Research | Sioux City, Iowa |
| Brandon Essink | Meridian Clinical Research | Omaha, Nebraska |
| Carlos Fierro | Johnson County Clin-Trials | Lenexa, Kansas |
| Veronica Fragoso | DM Clinical Research - Texas Center for Drug Development | Houston, Texas |
| Carl Griffin | Lynn Health Science Institute | Oklahoma City, Oklahoma |
| Greg Hachigian | Benchmark Research | Sacramento, California |
| Shishir Khetan | Meridian Clinical Research | Rockville, Maryland |
| Michael Koren | Jacksonville Center for Clinical Research | Jacksonville, Florida |
| Vicki Miller | DM Clinical Research | Tomball, Texas |
| Paul Pickrell | Tekton Research Inc. | Austin, Texas |
| Rachel Presti | Infectious Disease Clinical Research | Saint Louis, Missouri |
| Howard Schwartz | Research Centers of America | Hollywood, Florida |
| William Seger | Benchmark Research | Fort Worth, Texas |
| Charles Harper. Keith Vrbicky, | Meridian Clinical Research | Norfolk, Nebraska |
| Larkin Wadsworth | Sundance Clinical Research | Saint Louis, Missouri |
| Stephen Walsh | Brigham and Womens Hospital | Boston, Massachusetts |
| Jordan Whatley | Meridian Clinical Research | Baton Rouge, Louisiana |
| Barton Williams | Trial Management Associates LLC | Wilmington, North Carolina |
| Marcus Zervos | Henry Ford Health System | Detroit, Michigan |

### SUPPLEMENTARY METHODS

#### Study Design

This is an open-label, ongoing, 6-part, phase 2/3 study to evaluate the immunogenicity, safety, and reactogenicity of various modified mRNA-1273 vaccine candidates against Covid-19 administered as boosters (mRNA1273.211, mRNA1273, mRNA1273.617.2, mRNA-1273.213, mRNA1273.529, and mRNA-1273.214; clintrials.gov NCT04927065). Part G interim results are reported here. In part G, the bivalent mRNA-1273.214 vaccine which contains 25 ug each of two mRNAs encoding the ancestral SARS-CoV-2 and omicron variant (B.1.1.529) spike sequences is tested as a second booster dose at 50-µg in comparison to mRNA-1273 given as a second booster dose in part F, cohort 2) of the study. Study part F Cohort 2 will assess whether a single booster dose of the mRNA-1273.529 administered as a second booster dose elicits a similar or superior antibody response to the omicron (B.1.1.529) compared to a single booster dose of 50-µg mRNA-1273 as a second booster dose (part F, cohort 2, 50-µg mRNA-1273).

Enrollment of the mRNA-1273.214 50-µg second boost arm is initiated upon completion of enrollment of the mRNA-1273 50-µg arm in cohort 2 of part F. Part G evaluates the immunogenicity, safety, and reactogenicity of 50-µg of the mRNA1273.214 vaccine candidate when administered as a second booster dose to adults who have previously received 2 doses of 100 µg mRNA-1273 as a primary series and a first booster dose of mRNA-1273 (50 µg) in the Coronavirus Efficacy (COVE) trial or under US emergency use authorization (EUA). The immunogenicity of mRNA-1273.214 50-µg is compared to that induced after the second booster dose of mRNA 1273 (Part F, cohort 2, 50-µg mRNA-1273).

#### Part G Study Eligibility Criteria

##### Inclusion Criteria:

Each participant must meet all of the following criteria to be enrolled in this study:

1. Male or female, at least 18 years of age at the time of consent (Screening Visit).
2. Investigator's assessment that participant understands and is willing and physically able to comply with protocol-mandated follow-up, including all procedures.
3. Participant has provided written informed consent for participation in this study, including all evaluations and procedures as specified in this protocol.
4. Female participants of nonchildbearing potential may be enrolled in the study. Nonchildbearing potential is defined as surgically sterile (history of bilateral tubal ligation, bilateral oophorectomy, hysterectomy) or postmenopausal (defined as amenorrhea for ≥ 12 consecutive months prior to Screening [Day 0] without an alternative medical cause). A follicle-stimulating hormone level may be measured at the discretion of the investigator to confirm postmenopausal status.
5. Female participants of childbearing potential may be enrolled in the study if the participant fulfills all of the following criteria:
   - Has a negative pregnancy test on the day of vaccination (day 1).
   - Has practiced adequate contraception or has abstained from all activities that could result in pregnancy for at least 28 days prior to Day 1.
   - Has agreed to continue adequate contraception through 3 months following vaccination.
   - Is not currently breastfeeding.

Adequate female contraception is defined as consistent and correct use of a Food and Drug Administration approved contraceptive method in accordance with the product label.

6. Participant must have been either previously enrolled in the phase 3 mRNA 1273 COVE trial,[1,2] must have received 2 doses of mRNA 1273 in that study, with his/her second dose at least 6 months prior to enrollment in this study, and must be currently enrolled and compliant in that study (i.e., has not withdrawn or discontinued early); or participant must have received 2 doses of mRNA-1273 under the EUA with their second dose at least 6 months prior to enrollment in this study; or have received a 2 dose primary series of mRNA-1273 followed by a 50-µg booster dose of mRNA-1273 in the mRNA-1273 COVE trial or under EUA at least 3 months prior to enrollment this study; and able to provide proof of vaccination status at the time of screening (day 1).

**Exclusion Criteria:**

Participants meeting any of the following criteria at the Screening Visit, unless noted otherwise, will be excluded from the study:

1. Had significant exposure to someone with SARS-CoV-2 infection or coronavirus disease 2019 (Covid-19) in the past 14 days, as defined by the CDC as a close contact of someone who has Covid-19).
2. Has known history of SARS-CoV-2 infection within 3 months prior to enrollment.
3. Is acutely ill or febrile (temperature ≥38.0°C [100.4°F]) less than 72 hours prior to or at the Screening Visit or day 1. Participants meeting this criterion may be rescheduled and will retain their initially assigned participant number.
4. Currently has symptomatic acute or unstable chronic disease requiring medical or surgical care, to include significant change in therapy or hospitalization for worsening disease, at the discretion of the investigator.
5. Has a medical, psychiatric, or occupational condition that may pose additional risk as a result of participation, or that could interfere with safety assessments or interpretation of results according to the investigator’s judgment.
6. Has a current or previous diagnosis of immunocompromising condition to include human immunodeficiency virus, immune-mediated disease requiring immunosuppressive treatment, or other immunosuppressive condition.
7. Has received systemic immunosuppressants or immune-modifying drugs for >14 days in total within 6 months prior to Screening (for corticosteroids ≥10 mg/day of prednisone equivalent) or is anticipating the need for immunosuppressive treatment at any time during participation in the study.
8. Has known or suspected allergy or history of anaphylaxis, urticaria, or other significant AR to the vaccine or its excipients.
9. Has a documented history of myocarditis or pericarditis within 2 months prior to Screening Visit (day 0).
10. Coagulopathy or bleeding disorder considered a contraindication to intramuscular (IM) injection or phlebotomy.
11. Has received or plans to receive any licensed vaccine ≤ 28 days prior to the injection (day 1) or a licensed vaccine within 28 days before or after the study injection, with the exception of influenza vaccines, which may be given 14 days before or after receipt of a study vaccine.

12. Has received systemic immunoglobulins or blood products within 3 months prior to the Screening Visit (day 0) or plans for receipt during the study.
13. Has donated ≥450 mL of blood products within 28 days prior to the Screening Visit or plans to donate blood products during the study.
14. Plans to participate in an interventional clinical trial of an investigational vaccine or drug while participating in this study.
15. Is an immediate family member or household member of study personnel, study site staff, or Sponsor personnel.
16. Is currently experiencing an SAE in COVE trial at the time of screening for this study.

**Immunogenicity Objectives and Endpoints**

There were four pre-specified immunogenicity objectives at day 29 in the study (Table S1) including:

1) the second booster dose of 50-µg mRNA-1273.214 was non-inferior compared to the second booster dose of 50-µg mRNA-1273 based on the GMT ratio of mRNA-1273.214 against the omicron variant compared to mRNA-1273 against the omicron variant with a non-inferiority margin of 1.5
2) the second dose of 50-µg mRNA-1273.214 was non-inferior to the second booster dose of 50-µg mRNA-1273 against the omicron variant based on the difference in SRR with a non-inferiority margin of 10%
3) the second booster dose of 50-µg mRNA-1273.214 was non-inferior to the second booster dose of 50-µg mRNA-1273 against the ancestral SARS-CoV-2 (D614G) based on the GMT ratio of mRNA-1273.214 against the ancestral SARS-CoV-2 (D614G) compared to mRNA-1273 against the ancestral SARS-CoV-2 (D614G) with a non-inferiority margin of 1.5
4) the second booster dose of 50-µg mRNA-1273.214 was superior to the second booster dose of 50-µg mRNA-1273 against omicron B.1.1.529 based on the GMT ratio of mRNA-1273.214 against the omicron variant compared to mRNA-1273 against the omicron variant. The fourth objective is only tested if the first 3 non-inferiority hypotheses are demonstrated (Fig. S1)

If all primary immunogenicity hypotheses are demonstrated, the key secondary objective, the second booster dose of 50-µg mRNA-1273.214 was non-inferior to the booster dose of 50-µg of mRNA-1273 against ancestral SARS-CoV-2 (D614G) based on the difference in SRR at Day 29 with a non-inferiority margin of 10% is tested.

**Statistical Analysis of Immunogenicity**

In part G of the study, 50-µg mRNA-1273.214 as the second booster dose is compared to 50-µg mRNA-1273 as the second booster dose (active control arm in part F, Cohort 2). For the primary objective on immune response, 8 hypotheses are specified to be evaluated at days 29 and 91. Only interim analysis results for the day 29 hypotheses are presented in this report; hypotheses for day 91 will be tested later when day 91 data become available.

Below are the 4 hypotheses for day 29, day 91 hypotheses are provided in the online protocol and SAP.

1) 50-µg mRNA-1273.214, as a second booster dose, against the omicron (B.1.1.529) variant is non-inferior to the second booster dose of (50-µg) mRNA-1273 against the omicron (B.1.1.529) based on the GMT ratio of mRNA1273.214 compared to mRNA-1273 against omicron (B.1.1.529) at day 29 with a non-inferiority margin of 1.5.

2) 50-µg mRNA-1273.214, as a second booster dose, against the omicron (B.1.1.529) variant is non-inferior to the second booster dose of (50-µg) mRNA-1273 against omicron (B.1.1.529) based on the difference in SRR at day 29 with a non-inferiority margin of 10%.
3) 50-µg mRNA-1273.214, as a second booster dose, against ancestral SARS-CoV-2 (D614G) is non-inferior to the second booster dose of (50-µg) mRNA-1273 against ancestral SARS-CoV-2 (D614G) based on the GMT ratio of mRNA-1273.214 compared to mRNA-1273 against the ancestral SARS-CoV-2 (D614G) at day 29 with a non-inferiority margin of 1.5.
4) 50-µg mRNA-1273.214, as a second booster dose, against the omicron (B.1.1.529) variant is superior to the second booster dose of (50-µg) mRNA-1273 against B.1.1.529 based on the GMT ratio of mRNA-1273.214 compared to mRNA-1273 against the omicron (B.1.1.529) at day 29.

For the primary immunogenicity objective, an alpha of 0.05 (two-sided) is allocated to the two time points (days 29 and 91). Days 29 and 91 each have an alpha of 0.025 (two-sided) for hypotheses testing. The primary immunogenicity objective is considered met if non-inferiority against omicron B.1.1.529 based on GMR and SRR difference, and non-inferiority against ancestral SARS-CoV-2 (D614G) based on GMR are demonstrated at days 29 or 91.

For the key secondary immunogenicity objective, there are 2 hypotheses to be tested (days 29 and 91 will each have alpha of 0.025 [two-sided] for hypotheses testing):

1) 50-µg mRNA-1273.214, as a second booster dose, against ancestral SARS-CoV-2 (D614G) is non-inferior to the booster dose of (50-µg) mRNA-1273 against ancestral SARS-CoV-2 (D614G) based on the difference in SRR at Day 29 with a non-inferiority margin of 10%.
2) 50-µg mRNA-1273.214, as a second booster dose, against ancestral SARSCoV2 (D614G) is non-inferior to the booster dose of (50-µg) mRNA1273 against ancestral SARS-CoV-2 (D614G) based on the difference in SRR at Day 91 with a non-inferiority margin of 10%.

The planned target enrollments are approximately 375 participants for 50-µg mRNA-1273.214 in part G and for the comparator 50-µg mRNA-1273 in part F (cohort 2). Hypotheses testing is performed at days 29 and 91, alpha of 0.025 (2-sided) will be allocated equally to each one of the two time points. Assuming 20% of participants will be excluded from the PP Set for Immunogenicity – SARS-CoV-2 negative, with approximately 300 participants in 50-µg mRNA-1273.214 and 300 participants in 50-µg mRNA-1273 (part F, Cohort 2, 50-µg mRNA-1273) in the PP Set for Immunogenicity and SARS-CoV-2 negative, there is approximately 71% global power to demonstrate the primary immunogenicity objectives with alpha of 0.025 (2-sided) at each time point. The assumptions are: the true GMR (mRNA-1273.214 second booster vs 50-µg mRNA-1273 second booster) against the omicron variant (B.1.1.529) is 1.5, the true GMR against ancestral SARS-CoV-2 (D614G) is 1, the standard deviation of the log-transformed titer is 1.5, and the non-inferiority margin for GMR is 1.5, the true SRR against B.1.1.529 after mRNA-1273.214 as a second booster dose is 90% (same assumption for both 50-µg mRNA-1273.214 and 50-µg mRNA-1273), and non-inferiority margin for SRR difference is 10%. With approximately 375 participants exposed to 50-µg mRNA-1273.214, there is at least 90% probability in this group to observe 1 participant reporting an AE if the true rate of AEs is 1%.

An analysis of covariance (ANCOVA) model was performed to assess the difference in antibody responses between mRNA-1273.214 and mRNA-1273 booster doses, with antibody titers post-booster as a dependent variable, and a group variable (mRNA-1273.214 and mRNA-1273) as the fixed effect, adjusting for age groups (<65, ≥65 years) and pre-booster antibody titers. The GMTs (95% CI) estimated by the geometric least square mean (GLSM) from the model for each group and the GMR (mRNA-1273.214 compared with mRNA-1273) estimated by the ratio of GLSM from the model (97.5% CIs) are provided. The 97.5% CI for GMR was used to assess the between group difference in antibody responses.

Seroresponse is defined as ≥4 × LLOQ for those with a pre-dose 1 of a primary series baseline < LLOQ; ≥ 4-foldrise for those with pre-dose 1 of primary series baseline ≥ LLOQ. For participants without pre-dose 1 antibody titer information, seroresponse is defined as ≥4X LLOQ for those with negative SARS-CoV-2 status at pre-dose 1 of the primary series, and antibody titers are imputed as <LLOQ at pre-dose 1 of primary series. For participants who are without SARS-CoV-2 status information at pre-dose 1 of primary series, their pre-booster SARS-CoV-2 status is used to impute their SARS-CoV-2 status at their pre-dose 1 of primary series.

The SRR of each arm against ancestral SARS-CoV-2 (D614G) and variants, defined as the percentage of participants achieving SRR against ancestral SARS-CoV-2 (D614G) and variants respectively, are provided for each arm with the 95% CI calculated using the Clopper Pearson method. The differences of SRR between mRNA-1273.214 and mRNA-1273 are calculated with 97.5% CI based on stratified Miettinen-Nurminen method adjusting for age groups.

**Immunogenicity Assays**

***SARS-CoV-2 Spike-Pseudotyped Virus Neutralization Assay***

SARS-CoV-2 neutralizing antibodies (nAb) in samples were assessed using the validated SARS-CoV-2 Spike (S)-Pseudotyped Virus Neutralization Assay (PsVNA) in 293/ACE2 cells.[3] The PsVNA quantifies nAb using lentivirus particles that express ancestral SARS-CoV-2 Wuhan-Hu-1 full-length spike proteins with the following amino acid substitutions (prototype [D614G]; omicron [B.1.1.529; BA.1] with the following amino acid changes in the spike protein [A67V, ΔH69-V70, T95I, G142D, Δ143-145, Δ211/L212II, ins214EPE, G339D, S371L, S373P, S375F, K417N, N440K, G446S, S477N, T478K, E484A, Q493R, G496S, Q498R, N501Y, Y505H, T547K, D614G, H655Y, N679K, P681H, N764K, D796Y, N856K, Q954H, N969K, and L981F]; beta (B.1.351 [501Y-V2] L18F, D80A, D215G, Δ242-244, R246I, K417N, E484K, N501Y, D614G, and A701V); and delta ([B.1.617.2; AY.3] T19R, G142D, Δ156-157, R158G, L452R, T478K, D614G, P681R, D950N)]) on their surface, and contain a firefly luciferase reporter gene for quantitative measurements of infection in transduced 293T cells expressing high levels of ACE2 (293T/ACE2 cells) by relative luminescence units (RLU). Serial dilution of antibodies was used to produce a dose-response curve. Neutralization was measured as the serum dilution at which RLU was reduced by 50% (ID50) relative to mean RLU in virus control wells (cells + virus but no sample) after subtraction of mean RLU in cell control wells (cells only). Positive controls were included on each assay plate in order to follow stability over time. Omicron BA.4/BA.5 was assessed using a research grade pseudovirus lentivirus neutralization assay.

***SARS-CoV-2 Meso Scale Discovery (MSD) assay***

The validated Meso Scale Discovery (MSD, Rockville, MD) assay (SARSCOV2S2P [VAC123]; https://www.mesoscale.com/products/v-plex-sars-cov-2-panel-24-igg-kit-k15575u/) uses an indirect, quantitative, electrochemiluminescence method to detect SARS-CoV-2 binding IgG antibodies that bind to the SARS-CoV-2 full-length spike protein (Wuhan-Hu-1 ancestral SARS-CoV-2; beta [B.1.351] with the following amino acid changes in the spike protein [L18F, D80A,

D215G, Δ242-244, R246I, K417N, E484K, N501Y, D614G, and A701V]; alpha [B.1.1.7] with the following amino acid changes in the spike protein [ΔH69-V70, ΔY144, N501Y, A570D, D614G, P681H, T761I, S982A, and D1118H]; gamma [P.1] with the following amino acid changes in the spike protein [L18F, T20N, P26S, D138Y, R190S, K417T, E484K, N501Y, D614G, H655Y, T1027I, and V1176F]); delta [B.1.617.2; AY.4; Alt Seq 2] with the following amino acid changes in the spike protein [T19R, T95I, G142D, Δ156/157, R158G, L452R, T478K, D614G, P681R, and D950N]); omicron [B.1.1.529; BA.1] with the following amino acid changes in the spike protein [A67V, ΔH69-V70, T95I, G142D, Δ143-145, Δ211/L212I, ins214EPE, G339D, S371L, S373P, S375F, K417N, N440K, G446S, S477N, T478K, E484A, Q493R, G496S, Q498R, N501Y, Y505H, T547K, D614G, H655Y, N679K, P681H, N764K, D796Y, N856K, Q954H, N969K, and L981F]) in human serum. The assay was performed by PPD, Wilmington, NC. The assay is based on MSD technology which employs capture molecule MULTI-SPOT® microtiter plates fitted with a series of electrodes.

**Incidence of SARS-CoV-2 Infection and Covid-19**

Vaccine efficacy is not formally assessed in this trial, but Covid-19 and SARS-CoV-2 infection were actively surveilled through weekly contact and blood draws. An exploratory objective of the study is to assess symptomatic and asymptomatic SARS-CoV-2 infection. SARS-CoV-2 infection is a combination of symptomatic infection (Covid-19) and asymptomatic SARS-CoV-2 infection for participants with negative SARS-CoV-2 status pre-booster.

***SARS-CoV-2 infection***

SARS-CoV-2 infection is defined in participants with negative SARS-CoV-2 status pre-booster by either bAb levels against SARS-CoV-2 nucleocapsid protein negative (as measured by Roche Elecsys) at day 1 that becomes positive (as measured by Roche Elecsys) starting at day 29 or later, OR a positive RT-PCR counted starting 14 days after the booster dose.

For the analysis, documented infection is counted starting 14 days after the booster dose, which requires positive serology test result based on bAb specific to SARS-CoV-2 nucleocapsid at day 29 or later, or a positive RT-PCR result starting 14 days after the booster dose.

The date of documented infection is the earlier of the date of a positive post-baseline RT-PCR result or of positive serology test result based on bAb specific to SARS-CoV-2 nucleocapsid. The time to first SARS-CoV-2 infection is calculated as the date of the 1st documented infection minus the date of injection + 1. Cases are counted starting 14 days after the injection (date of documented infection minus date of the injection ≥14). SARS-CoV-2 infection cases are summarized based on tests performed at least 14 days after the booster dose.

***Asymptomatic SARS-CoV-2 Infection***

Asymptomatic SARS-CoV-2 infection is measured by RT-PCR of nasal swabs and/or serology tests obtained at post-baseline study visits counted starting 14 days after the injection in participants with negative SARS-CoV-2 status pre-booster. Asymptomatic SARS-CoV-2 infection is identified by the absence of symptoms and infections as detected by RT-PCR or serology tests. Specifically, the absence of Covid-19 symptoms AND at least either a positive serology test result based on bAb specific to SARS-CoV-2 nucleocapsid protein day 29 or later, when blood samples for immunogenicity are collected, or a positive RT-PCR test at scheduled or unscheduled/illness visits. The date of documented asymptomatic infection is the earlier date of positive serology test result based on bAb specific to SARS-CoV-2 nucleocapsid due to infection, or positive RT-PCR, with absence of symptoms. The time to the asymptomatic SARS-CoV-2 infection is calculated as the date of asymptomatic SARS-CoV-2 infection minus the date of injection + 1.

***Symptomatic SARS-CoV-2 Infection (Covid-19)***

Symptomatic SARS-CoV-2 Infection (Covid-19) is defined as the incidence of the first occurrence of symptomatic SARS-CoV-2 infection measured by RT-PCR of nasal swabs starting 14 days after the booster dose. Surveillance for Covid-19 symptoms is conducted via weekly contact and blood draw, and an illness visit to collect a nasopharyngeal swab was arranged for participants reporting Covid-19 symptoms.

Two definitions of symptomatic SARS-CoV-2 infection (Covid-19). This includes the primary case definition in the COVE trial[1,2] based on a positive post-baseline RT-PCR result AND at least TWO systemic symptoms (fever (≥ 38ºC/≥ 100.4ºF), chills, muscle and/or body aches [not related to exercise], headache, sore throat, new loss of taste/smell; OR at least ONE of respiratory signs/symptoms (cough, shortness of breath and/or difficulty breathing, OR clinical or radiographical evidence of pneumonia). The second case definition is based on CDC criteria for symptomatic disease defined as a positive post-baseline RT-PCR test AND at least ONE systemic or respiratory symptoms (fever [≥ 38ºC/≥100.4ºF], chills, cough, shortness of breath and/or difficulty breathing, fatigue, muscle and/or body aches [not related to exercise], headache, new loss of taste/smell, sore throat, congestion, runny nose, nausea, vomiting, or diarrhea).[4]

The date of a documented Covid-19 case is the later date of a symptom and the date of positive RT-PCR test, and the two dates should be within 14 days of each other. The time to the first occurrence of Covid-19 is calculated as the date of documented Covid-19 minus the date of injection + 1. Cases are counted starting 14 days after the injection (date of documented Covid-19 minus date of the injection ≥14).

**Figure S1: Statistical Testing Sequence**

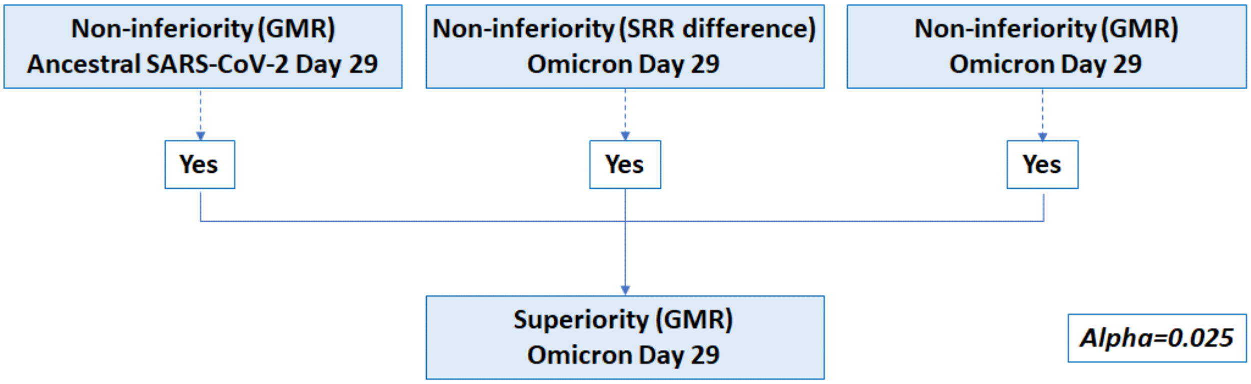


**Figure S1. Statistical Hypotheses Testing Strategy.** For Part G of the study, there were 4 corresponding clinical endpoints for the pre-specified primary objectives: (1) non-inferiority of the antibody response of the second booster dose of 50-µg mRNA-1273.214 compared with the second booster dose of 50-µg mRNA-1273 based on the GMR against omicron; 2) non-inferiority of the antibody response of the second dose of 50-µg mRNA-1273.214 compared to the second booster dose of 50-µg mRNA-1273 against omicron based on the difference in SRR; 3) non-inferiority of the antibody response of the second booster dose of 50-µg mRNA-1273.214 compared to the second booster dose of 50-µg mRNA-1273 based on GMR against the ancestral SARS-CoV-2 (D614G) and, 4) superiority of the antibody response of the second booster dose of 50-µg mRNA-1273.214 compared to the second booster dose of 50-µg mRNA-1273 based on the GMR against omicron (detailed in supplementary statistical methods). The endpoint testing sequence pre-specified that all 3 endpoints must first be met to test for the fourth endpoint. The immunogenicity endpoints are tested with an alpha of 0.025 (2-sided) for the interim analysis at 28 days after the booster dose (day 29). Non-inferiority is considered met when the lower bound of the 97.5% confidence interval (CI) of GMR is ≥0.67 and when SRR difference >-10%. Superiority is considered met when the lower bound of the 97.5% CI of GMR is >1. Superiority of the mRNA-1273.214 antibody response against omicron, compared to mRNA-1273, is considered demonstrated if superiority based on GMR is met at day 29. If non-inferiority is demonstrated for both omicron (based on GMR and SRR) and ancestral SARS-CoV-2 (D614G) (based on GMR), the lower bound of 97.5% CI of GMR is compared to 1, and if greater than 1, then superiority against omicron is demonstrated.

**Figure S2. Observed Neutralizing Antibody Titers Against SARS-CoV-2 (D614G) and Omicron after 50-μg of mRNA-1273.214 and mRNA-1273 Administered as Second Booster Doses**

A. Ancestral SARS-CoV-2 (D614G)

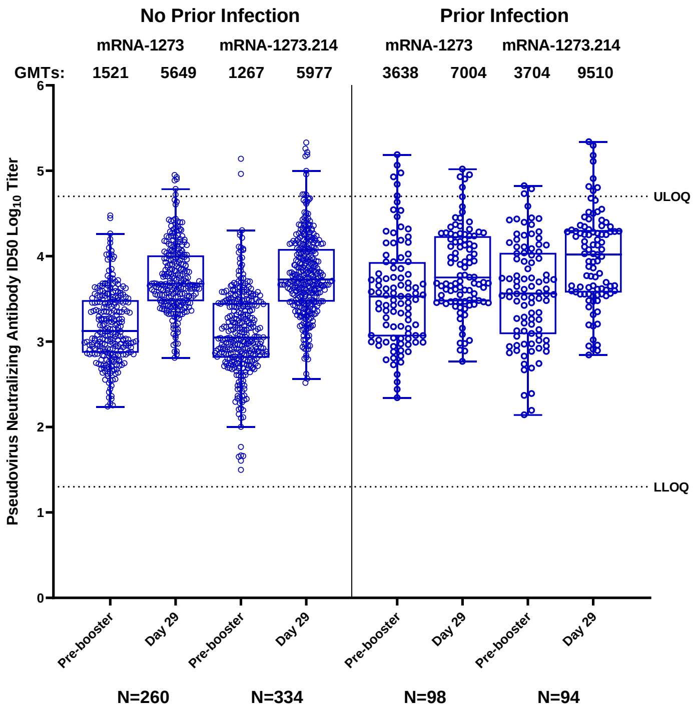


B. Omicron

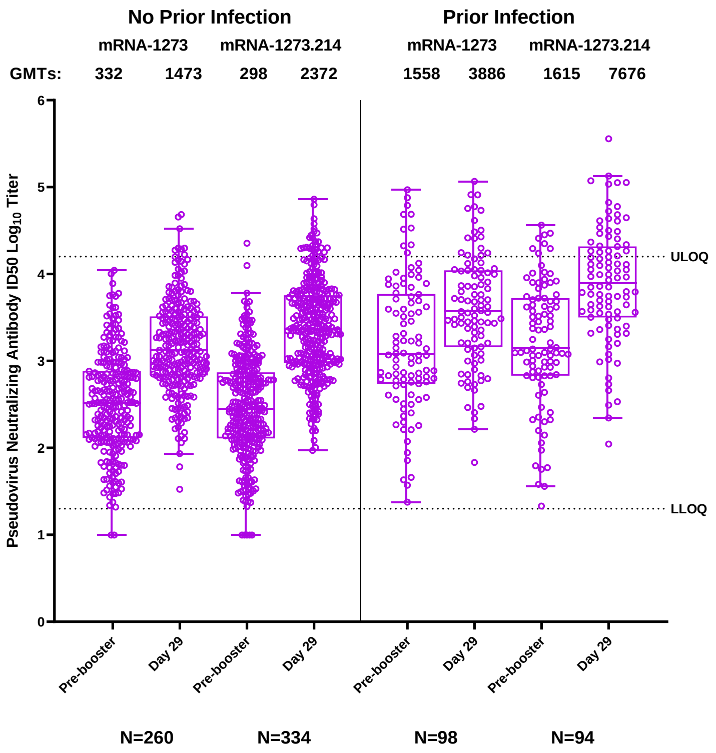

**Figure S2. Observed Neutralizing Antibody Titers Against SARS-CoV-2 (D614G) and Omicron after 50-µg of mRNA-1273.214 and mRNA-1273 Administered as Second Booster Doses.** The neutralizing antibody titers (ID50 $log_{10}$) in the pseudovirus assay against the ancestral SARS-CoV-2 (D614G) (Panel A; blue) or the omicron variant (Panel B; purple) are shown for serum samples collected before the second booster dose of 50-µg of mRNA-1273 or 50-µg of mRNA-1273.214 (pre-booster), and at 28 days after the second booster dose (day 29). The circles are the values from individual serum samples. Pseudovirus neutralizing antibody assay lower limits of quantification (LLOQ) are 18.5 for ancestral SARS-CoV-2 [D614G] and 19.9 for omicron; upper limits of quantification (ULOQ) are 45,118 for ancestral SARS-CoV-2 [D614G] and 15,502.7 for omicron. Corresponding $log_{10}$ values for LLOQs for the pseudovirus neutralizing antibody assay are 1.3 for both ancestral [D614G] and omicron, and ULOQs are 4.7 for ancestral SARS-CoV-2 [D614G] and 4.2 omicron. Boxes and horizontal bars denote interquartile (IQR) ranges and median endpoint titers; whisker endpoints are the maximum (75% percentile) and minimum (25% percentile) values below or above the median ±1.5 times the IQR. Refer to Table 2 for the observed geometric mean titers and geometric mean fold rises, and the estimated neutralizing antibody geometric mean titers and geometric mean ratios (ANCOVA model-based) following the second booster doses of 50-µg mRNA-1273 or 50-µg of mRNA-1273.214.

**Figure S3. Observed Neutralizing Antibody Titers Against Omicron BA.4 and BA.5 Subvariants after 50-µg of mRNA-1273.214 Administered as a Second Booster Dose**

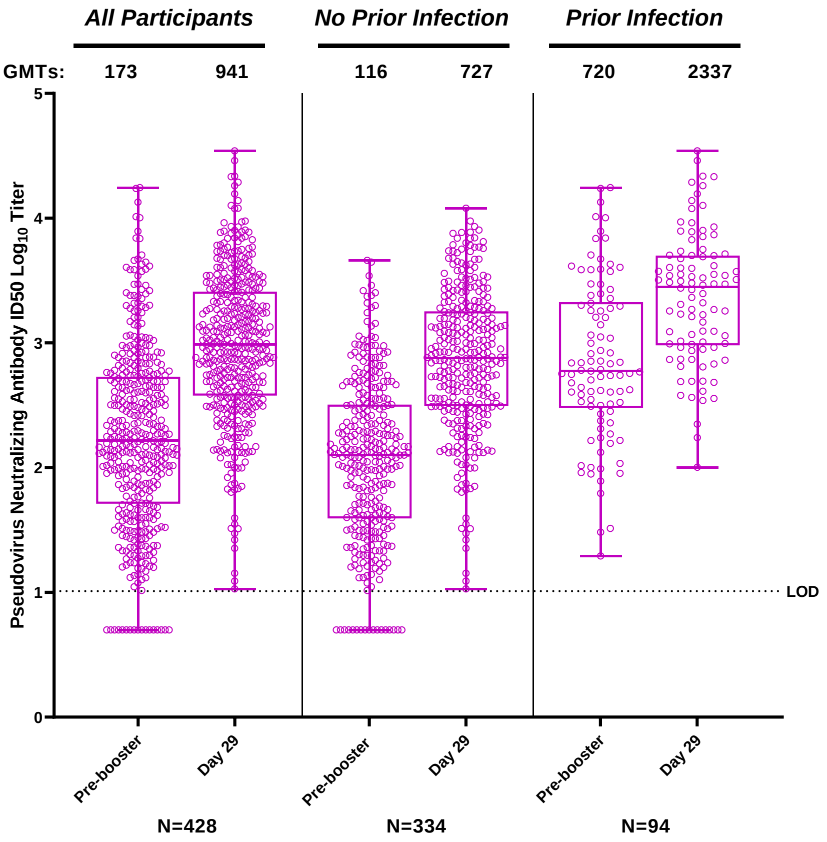


**Figure S3. Observed Neutralizing Antibody Titers Against Omicron BA.4 and BA.5 Subvariants after 50-µg of mRNA-1273.214 Administered as a Second Booster Dose.** The neutralizing antibody titers (ID50 $\log_{10}$) in the research grade pseudovirus assay against the omicron BA.4/BA.5 variants are shown for serum samples collected before the second booster dose of mRNA-1273.214 (pre-booster), and at 28 days after the second booster dose (day 29). Assay limit of detection (LOD) is 10 ($\log_{10}$=1). The circles are the values from individual serum samples. Boxes and horizontal bars denote interquartile (IQR) ranges and median endpoint titers; whisker endpoints are the maximum (75% percentile) and minimum (25% percentile) values below or above the median ±1.5 times the IQR. Observed GMTs are provided in Table S8.

**Figure S4. Observed Binding Antibody Levels Against Variants After 50-µg of mRNA-1273.214 and mRNA-1273 Administered as Second Booster Doses in All Participants**

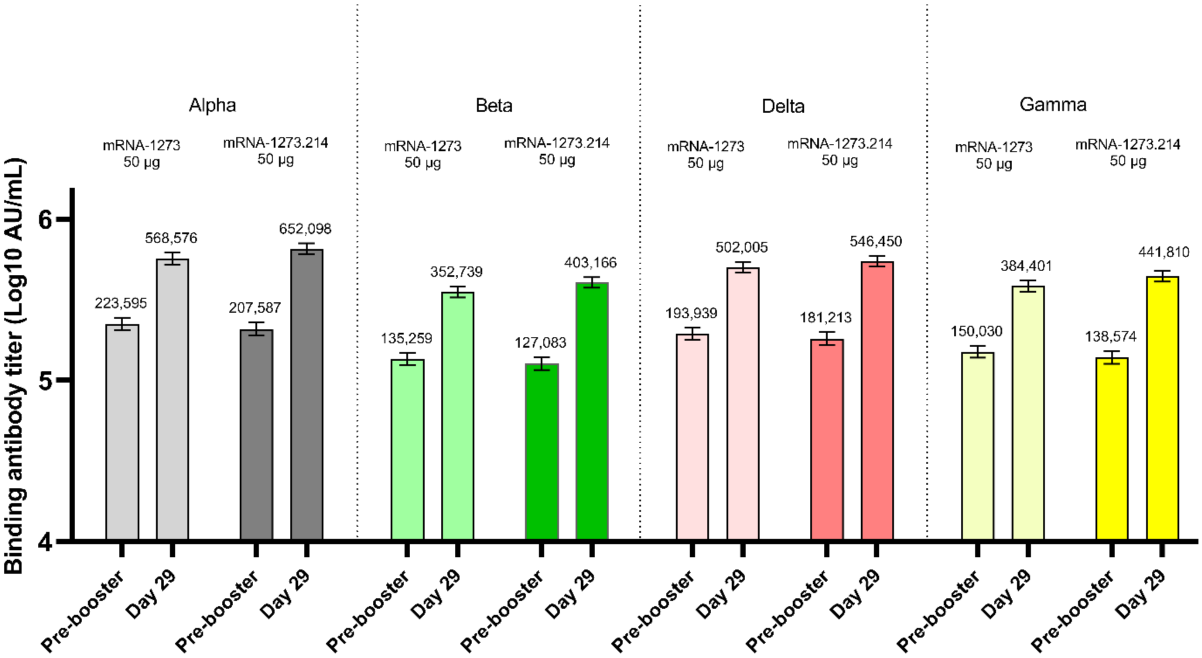


**Figure S4**. **Observed Binding Antibody Levels Against Variants After 50-µg of mRNA-1273.214 and mRNA-1273 Administered as Second Booster Doses in All Participants.** CI=confidence interval. The levels (AU/mL) of antibodies that specifically bind to the alpha, beta, delta, or gamma spike proteins were determined by the Mesoscale Discovery (MSD) Multiplex (VAC123) assay. Serum samples were collected before the second booster dose of 50-µg of mRNA-1273 or 50-µg of mRNA-1273.214 (pre-booster), and at 28 days after the second booster injection (day 29). Number of participants (n) ranges from 350 to 365 in mRNA-1273 group, and n ranges from 398 to 423 in mRNA-1273.214 group. Antibody values reported as below the lower limit of quantification (LLOQ) were 52 for alpha, 111 for beta, 150 for delta, 143 for gamma and were replaced by 0.5 times the LLOQ. Antibody values reported as greater than the upper limit of quantification (ULLQ) were 8,800,000 for alpha, 5,000,000 for beta, 8,000,000 for delta, 5,800,000 for gamma and were converted to the ULOQ if the actual values were not available. The 95% confidence intervals were calculated based on the t-distribution of the log-transformed values or the difference in the log-transformed values for the geometric mean value then back transformed to the original scale for presentation. Observed binding antibody titers are summarized in table S10.

**Table S1. Objectives and Endpoints for Part G Second Booster Doses of 50-µg mRNA-1273.214 in Participants Who Received 100 µg mRNA-1273 Primary Series and a Booster Dose of 50 µg mRNA-1273**

| Objectives | Endpoints |
|---|---|
| **Primary** | |
| • To demonstrate non-inferiority of the antibody response of a second booster dose of mRNA-1273.214 compared to mRNA-1273 (50-µg) when administered as a second booster dose against the omicron variant (B.1.1.529) at Day 29 or Day 91 based on GMT ratio and SRR difference at Day 29 or Day 91<br>• To demonstrate superiority of the antibody response of a second booster dose of mRNA-1273.214 compared to mRNA-1273 (50-µg) administered as a second booster dose against the omicron variant (B.1.1.529) based on GMT ratio at Day 29 or Day 91<br>• To demonstrate non-inferiority of the antibody response of a second booster dose of mRNA-1273.214 compared to mRNA-1273 (50-µg) when administered as a second booster dose against ancestral SARS-CoV-2 (D614G) based on GMT ratio at Day 29 or Day 91 | • GMT ratio of omicron-specific GMT of mRNA-1273.214 over the omicron-specific GMT of mRNA-1273 (Part F, Cohort 2, 50 µg mRNA-1273) at Day 29 and Day 91<br>• SRR difference between mRNA-1273.214 against omicron variant and mRNA-1273 against omicron variant at Day 29 and Day 91<br>• GMT ratio of ancestral SARS-CoV-2 (D614G) GMT of mRNA-1273.214 over ancestral SARS-CoV-2 (D614G) GMT of mRNA-1273 (Part F, Cohort 2, 50 µg mRNA-1273) at Day 29 or Day 91 |
| • To evaluate the safety and reactogenicity of mRNA-1273.214 | • Solicited local and systemic reactogenicity ARs during a 7-day follow-up period after vaccination<br>• Unsolicited AEs during the 28-day follow-up period after vaccination<br>• Serious AEs (SAEs), MAAEs, AEs leading to withdrawal and AESIs from Day 1 to EoS |
| **Key Secondary** | |
| • To demonstrate non-inferiority based on the SRR against ancestral SARS-CoV-2 (D614G) of a second booster dose of mRNA-1273.214 compared to a second booster dose of mRNA-1273 (50-µg) at Day 29 or Day 91 | • SRR difference between mRNA-1273.214 against ancestral SARS-CoV-2 (D614G) and mRNA-1273 against ancestral SARS-CoV-2 (D614G) at Day 29 and Day 91 |
| **Secondary** | |
| • To evaluate the immunogenicity of mRNA-1273.214 booster compared to mRNA-1273 booster administered as a | • GMT ratio of mRNA-1273.214 and mRNA-1273 against the omicron variant at all timepoints post-boost |

| Objectives | Endpoints |
|---|---|
| second booster dose at all timepoints post-boost | • SRR difference between mRNA-1273.214 against the omicron variant and mRNA-1273 against the omicron variant at all timepoints post-boost<br>• GMT ratio of mRNA-1273.214 and mRNA-1273 against ancestral SARS-CoV-2 (D614G) and other variants at all timepoints post-boost<br>• SRR difference between mRNA-1273.214 against ancestral SARS-CoV-2 (D614G) and other variants and mRNA-1273 against ancestral SARS-CoV-2 (D614G) and other variants at all timepoints post-boost |
| • To compare the immune response of mRNA-1273.214 as a second dose against the omicron variant compared to the priming series of mRNA-1273 | • GMT ratio and SRR difference of mRNA-1273.214 as a second booster dose against the omicron variant compared to the priming series of mRNA-1273 against ancestral SARS-CoV-2 (D614G) (historical control group) |
| **Exploratory** | |
| • To assess for symptomatic and asymptomatic SARS-CoV-2 infection | • Laboratory-confirmed symptomatic or asymptomatic SARS-CoV-2 infection will be defined in participants:<br>– Primary case definition per the (COVE) study<br>– Secondary case definition based on the CDC criteria: the presence of one of the CDC-listed symptoms (https://www.cdc.gov/coronavirus/2019-ncov/symptoms-testing/symptoms.html) and a positive reverse transcriptase polymerase chain reaction (RT-PCR) test on a respiratory sample<br>• Asymptomatic SARS-CoV-2 infection is defined as a positive RT-PCR test on a respiratory sample in the absence of symptoms or a positive serologic test for anti-nucleocapsid antibody after a negative test at time of enrollment |
| • To evaluate the genetic and/or phenotypic relationships of isolated SARS-CoV-2 strains to the vaccine sequence | • Characterize the SARS-CoV-2 genomic sequence of viral isolates and compare with the vaccine sequence<br>• Characterize the immune responses to vaccine breakthrough isolates |
| • To characterize the cellular immune response of mRNA-1273.214 as a booster against SARS-CoV-2 and other variants | • T-cell and B-cell response after the mRNA-1273.214 booster |

**Table S2. Analysis Sets**

| Set | Description |
|---|---|
| Full Analysis Set (FAS) | The FAS consists of all participants who receive investigational product (IP). |
| Per-Protocol (PP) Set for Immunogenicity | The PP Set for Immunogenicity consists of all participants in the FAS who received the planned dose of study vaccination and no major protocol deviations that impact key or critical data.<br>The PP Set will be used as the primary analysis set for analyses of immunogenicity for immunobridging. |
| PP Set for Immunogenicity – SARS-CoV-2 negative (PPSI-Neg) | Participants in the PPSI who have no serologic or virologic evidence of SARS-CoV-2 infection at baseline, i.e., who are SARS-CoV-2 negative, defined by both negative RT-PCR test for SARS-CoV-2 and negative serology test based on bAb specific to SARS-CoV-2 nucleocapsid<br>PPSI-Neg will be the primary analysis set for analyses of immunogenicity for between booster comparisons. |
| Solicited Safety Set | The Solicited Safety Set consists of all participants who receive IP and contribute any solicited AR data.<br>The Solicited Safety Set will be used for the analyses of solicited ARs. Participants will be included in the study arm corresponding to the dose of IP that they actually received. |
| Safety Set | The Safety Set consists of all participants who receive IP. The Safety Set will be used for all analyses of safety except for the solicited ARs. Participants will be included in the study arm corresponding to the dose of IP that they actually received. |
| Per-Protocol Set for Efficacy | The PP Set for Efficacy consists of all participants in the FAS who receive the planned dose of study vaccination, who are SARS-CoV-2 negative at baseline (ie, have a negative RT-PCR test for SARS-CoV-2 and a negative serology test based on bAb specific to SARS-CoV-2 nucleocapsid at baseline), and have no major protocol deviations that impact key or critical data. |

**Table S3. Solicited Local and Systemic Adverse Reactions Within 7 Days Following the Second Booster Injections of mRNA-1273.214 (50 µg) and mRNA-1273 (50 µg)**

| Adverse Reaction, N (%) | mRNA-1273.214 Second Booster Dose (50 µg) N=437 | mRNA-1273 Second Booster Dose (50 µg) N=351 |
|---|---|---|
| Solicited AR, N1 | 437 | 351 |
| Any Solicited AR | 380 (87.0) | 301 (85.8) |
| Grade 1 | 220 (50.3) | 184 (52.4) |
| Grade 2 | 125 (28.6) | 89 (25.4) |
| Grade 3 | 35 (8.0) | 28 (8.0) |
| Grade 4 | 0 | 0 |
| Any Solicited Local AR, N1 | 437 | 351 |
| Any Solicited Local AR | 347 (79.4) | 279 (79.5) |
| Grade 1 | 291 (66.6) | 239 (68.1) |
| Grade 2 | 41 (9.4) | 28 (8.0) |
| Grade 3 | 15 (3.4) | 12 (3.4) |
| Local AR, Pain, N1 | 437 | 351 |
| Pain | 338 (77.3) | 269 (76.6) |
| Grade 1 | 303 (69.3) | 241 (68.7) |
| Grade 2 | 31 (7.1) | 24 (6.8) |
| Grade 3 | 4 (0.9) | 4 (1.1) |
| Erythema, N1 | 437 | 351 |
| Erythema | 30 (6.9) | 13 (3.7) |
| Grade 1 | 15 (3.4) | 5 (1.4) |
| Grade 2 | 6 (1.4) | 6 (1.7) |
| Grade 3 | 9 (2.1) | 2 (0.6) |
| Swelling, N1 | 437 | 351 |
| Swelling | 30 (6.9) | 23 (6.6) |
| Grade 1 | 17 (3.9) | 13 (3.7) |
| Grade 2 | 8 (1.8) | 5 (1.4) |
| Grade 3 | 5 (1.1) | 5 (1.4) |
| Axillary Swelling or Tenderness, N1 | 437 | 351 |
| Axillary Swelling or Tenderness | 76 (17.4) | 54 (15.4) |
| Grade 1 | 71 (16.2) | 46 (13.1) |
| Grade 2 | 4 (0.9) | 4 (1.1) |
| Grade 3 | 1 (0.2) | 4 (1.1) |
| Any Systemic AR, N1 | 437 | 351 |
| Any Systemic AR | 307 (70.3) | 232 (66.1) |
| Grade 1 | 167 (38.2) | 124 (35.3) |
| Grade 2 | 116 (26.5) | 92 (26.2) |
| Grade 3 | 24 (5.5) | 16 (4.6) |
| Fever, N1 | 436 | 351 |

| | | |
|---|---|---|
| Fever | 19 (4.4) | 12 (3.4) |
| Grade 1 | 14 (3.2) | 9 (2.6) |
| Grade 2 | 4 (0.9) | 3 (0.9) |
| Grade 3 | 1 (0.2) | 0 |
| Headache, N1 | 437 | 350 |
| Headache | 192 (43.9) | 144 (41.1) |
| Grade 1 | 150 (34.3) | 112 (32.0) |
| Grade 2 | 37 (8.5) | 30 (8.6) |
| Grade 3 | 5 (1.1) | 2 (0.6) |
| Fatigue, N1 | 437 | 350 |
| Fatigue | 240 (54.9) | 180 (51.4) |
| Grade 1 | 125 (28.6) | 95 (27.1) |
| Grade 2 | 100 (22.9) | 74 (21.1) |
| Grade 3 | 15 (3.4) | 11 (3.1) |
| Myalgia, N1 | 437 | 350 |
| Myalgia | 173 (39.6) | 135 (38.6) |
| Grade 1 | 101 (23.1) | 68 (19.4) |
| Grade 2 | 62 (14.2) | 54 (15.4) |
| Grade 3 | 10 (2.3) | 13 (3.7) |
| Arthralgia, N1 | 437 | 350 |
| Arthralgia | 136 (31.1) | 111 (31.7) |
| Grade 1 | 93 (21.3) | 70 (20.0) |
| Grade 2 | 39 (8.9) | 38 (10.9) |
| Grade 3 | 4 (0.9) | 3 (0.9) |
| Nausea/ vomiting, N1 | 437 | 350 |
| Nausea / vomiting | 45 (10.3) | 35 (10.0) |
| Grade 1 | 39 (8.9) | 27 (7.7) |
| Grade 2 | 5 (1.1) | 8 (2.3) |
| Grade 3 | 1 (0.2) | 0 |
| Chills, N1 | 437 | 350 |
| Chills | 104 (23.8) | 74 (21.1) |
| Grade 1 | 65 (14.9) | 46 (13.1) |
| Grade 2 | 38 (8.7) | 27 (7.7) |
| Grade 3 | 1 (0.2) | 1 (0.3) |

AR=adverse reaction. Any=Grade 1 or higher. N1=Number of exposed participants with any information about the adverse event. Percentages are based on the number of participants who submitted any data for the event. Data cutoff date was April 27, 2022.

**Table S4. Summary of Unsolicited Adverse Events ≤28 Days Post-booster Dose**

| n (%) | mRNA-1273.214 50 µg (N=437) | mRNA-1273 50 µg (N=377) |
|---|---|---|
| **Unsolicited AEs Regardless of Relationship to Study Vaccination** | | |
| All | 81 (18.5) | 78 (20.7) |
| Serious | 2 (0.5) | 1 (0.3) |
| Fatal | 0 | 0 |
| Medically attended | 43 (9.8) | 52 (13.8) |
| Leading to discontinuation from study | 0 | 0 |
| Grade 3 or higher | 4 (0.9) | 3 (0.8) |
| **Unsolicited AEs related to study vaccination** | | |
| All | 25 (5.7) | 22 (5.8) |
| Serious | 0 | 0 |
| Fatal | 0 | 0 |
| Medically attended | 2 (0.5)* | 2 (0.5)* |
| Leading to discontinuation from study | 0 | 0 |
| Grade 3 or higher | 1 (0.2) | 2 (0.5) |

AE, adverse event. An adverse event was defined as any event not present prior to study vaccination or any event already present that worsened in intensity or frequency after vaccination. Percentages are based on the number of participants in the safety set. *Medically attended AEs of fatigue (grade 2) and dermatitis (grade 1) occurred in the mRNA-1273.214 group and of hypertension and urticaria (both grade 1) in the mRNA-1273 groups.

**Table S5. Geometric Mean Neutralizing Antibody Titer Ratio and Seroresponse Difference Against Ancestral SARS-CoV-2 (D614G) and Omicron after 50-µg of mRNA-1273.214 and mRNA-1273 Administered as Second Booster Doses in All Participants Regardless of Evidence of Prior SARS-CoV-2 Infection**

| | Ancestral SARS-CoV-2 (D614G) | | Omicron | |
|---|---|---|---|---|
| | 50 µg mRNA-1273.214 Booster Dose* | 50 ug mRNA-1273 Booster dose* | 50 µg mRNA-1273.214 Booster Dose* | 50 ug mRNA-1273 Booster dose* |
| | N=428 | N=367 | N=428 | N=367 |
| Pre-booster n† | 428 | 367 | 428 | 367 |
| Observed GMT (95% CI)§ | 1603.4 (1420.3-1810.0) | 1944.8 (1725.4-2192.1) | 432.1 (372.5-501.2) | 512.0 (433.4-604.8) |
| Day 29, n† | 428 | 367 | 428 | 367 |
| Observed GMT (95% CI)§ | 6619.0 (5941.7-7373.5) | 6047.5 (5465.9-6691.0) | 3070.4 (2685.4-3510.6) | 1932.8 (1681.2-2222.0) |
| GMFR (95% CI)§ | 4.1 (3.8-4.4) | 3.1 (2.9-3.4) | 7.1 (6.5-7.8) | 3.8 (3.4-4.2) |
| Estimated GMT (95% CI)¶ | 6555.7 (6122.3-7019.7) | 5301.4 (4931.8-5698.7) | 3232.5 (2951.8-3539.9) | 1815.1 (1650.0-1996.7) |
| GMR (97.5% CI)¶ǁ | 1.24 (1.12-1.37) | | 1.78 (1.56-2.04) | |
| Day 29 SRR, n/N1 %‡ | 383/383, 100 | 347/347, 100 | 380/380, 100 | 340/342, 99.4 |
| (95% CI) | (99.0-100) | (98.9-100) | (99.0-100) | (97.9-99.9) |
| Difference (97.5% CI)†† | 0 | | 1.2 (-1.3, 3.7) | |

ANCOVA=analysis of covariance, CI = Confidence interval, GMT=geometric mean titer. GMR = Geometric mean ratio, mRNA-1273.214 vs mRNA-1273. ID50, 50% inhibitory dose; LLOQ, lower limit of quantification; LS, least squares; ULOQ, upper limit of quantification. Antibody values assessed by pseudovirus neutralizing antibody assay reported as below the LLOQ (18.5 for ancestral [D614G] and 19.9 for omicron are replaced by 0.5 × LLOQ. Values greater than ULOQ (45,118) for ancestral [D614G] and (15,502.7) for omicron are replaced by the ULOQ if actual values are not available.

†Number of participants with non-missing data at the timepoint (baseline or post-baseline).

§95% CI based on the t-distribution of log-transformed values or difference in the log-transformed values for GMT value and GMT fold-rise, respectively, then back transformed to the original scale.

¶Log-transformed antibody levels are analyzed using an ANCOVA model with the treatment variable as fixed effect, adjusting for age group (<65, ≥65 years), pre-booster titers and prior SARS-CoV-2 infection. The resulting LS means, difference of LS means, and 95% CI are back transformed to the original scale.

ǁ97.5% CI was calculated by stratified Miettinen-Nurminen method adjusted by age group and prior SARS-CoV-2 infection.

‡Seroresponse at a participant level defined as a change from <LLOQ to ≥4 × LLOQ, or at least a 4-fold rise if baseline is ≥LLOQ; comparison to pre-vaccination baseline. Percentages were based on the number of participants with non-missing data at baseline and the corresponding time point. 95% CI calculated using the Clopper-Pearson method.

††The SRR difference is a calculated common risk difference using inverse-variance stratum weights and the middle point of Miettinen-Nurminen confidence limits of each one of the stratum risk differences. The stratified Miettinen-Nurminen estimate and the CI cannot be calculated when the seroresponse rate in both groups is 100%, absolute difference is reported.

**Table S6. Geometric Mean Neutralizing Antibody Titer Ratio and Seroresponse Difference Against Ancestral SARS-CoV-2 (D614G) and Omicron after 50-µg of mRNA-1273.214 and mRNA-1273 administered as Second Booster Doses in Participants with Evidence of Prior SARS-CoV-2 Infection**

| | **Ancestral SARS-CoV-2 (D614G)** | | **Omicron** | |
|---|---|---|---|---|
| | **50 µg mRNA-1273.214 Booster Dose*** | **50 ug mRNA-1273 Booster dose*** | **50 µg mRNA-1273.214 Booster Dose*** | **50 ug mRNA-1273 Booster dose*** |
| | **N=94** | **N=98** | **N=94** | **N=98** |
| Pre-booster n† | | | | |
| Observed GMT (95% CI)§ | 3704.0 (2793.2-4911.7) | 3638.0 (2742.0-4826.6) | 1614.6 (1149.7-2267.7) | 1558.4 (1088.9-2230.1) |
| Day 29, n† | | | | |
| Observed GMT (95% CI)§ | 9509.7 (7345.9-12,310.9) | 7003.5 (5592.6-8770.4) | 7676.2 (5618.2-10,488.1) | 3885.6 (2877.8-5246.4) |
| GMFR (95% CI)§ | 2.6 (2.2-2.9) | 1.9 (1.6-2.2) | 4.8 (4.0-5.7) | 2.5 (2.1-3.0) |
| Estimated GMT (95% CI)¶ | 9891.5 (8732.2-11204.8) | 7776.5 (6813.0-8876.3) | 7669.2 (6470.7-9089.6) | 4041.5 (3375.1-4839.5) |
| GMR (95.0% CI)¶‖ | 1.27 (1.07-1.51) | | 1.90 (1.50-2.40) | |
| Day 29 SRR, n/N1 %‡ (95% CI) | 49/49, 100 (92.7-100.0) | 79/79, 100 (95.4-100.0) | 47/47, 100 (92.5-100.0) | 76/76, 100 (95.3-100.0) |
| Difference (95.0% CI)†† | 0 | | 0 | |

ANCOVA=analysis of covariance, CI = Confidence interval, GMT=geometric mean titer. GMR = Geometric mean ratio, mRNA-1273.214 vs mRNA-1273. ID50, 50% inhibitory dose; LLOQ, lower limit of quantification; LS, least squares; ULOQ, upper limit of quantification. Antibody values assessed by pseudovirus neutralizing antibody assay reported as below the LLOQ (18.5 for ancestral SARS-CoV-2 [D614G] and 19.9 for omicron are replaced by 0.5 × LLOQ. Values greater than ULOQ (45,118) for ancestral SARS-CoV-2 [D614G] and (15,502.7) for omicron are replaced by the ULOQ if actual values are not available.
†Number of participants with non-missing data at the timepoint (baseline or post-baseline).
§95% CI based on the t-distribution of log-transformed values or difference in the log-transformed values for GMT value and GMT fold-rise, respectively, then back transformed to the original scale.
¶Log-transformed antibody levels are analyzed using an ANCOVA model with the treatment variable as fixed effect, adjusting for age group (<65, ≥65 years) and pre-booster titers. The resulting LS means, difference of LS means, and 95% CI are back transformed to the original scale for presentation.
‖95% CI was calculated by stratified Miettinen-Nurminen method adjusted by age group.
‡Seroresponse at a participant level defined as a change from <LLOQ to ≥4 × LLOQ, or at least a 4-fold rise if baseline is ≥LLOQ; comparison to pre-vaccination baseline. Percentages were based on the number of participants with non-missing data at baseline and the corresponding time point. 95% CI calculated using the Clopper-Pearson method.
††The SRR difference is a calculated common risk difference using inverse-variance stratum weights and the middle point of Miettinen-Nurminen confidence limits of each one of the stratum risk differences. The stratified Miettinen-Nurminen estimate and the CI cannot be calculated when the seroresponse rate in both groups is 100%, absolute difference is reported.

**Table S7. Observed Neutralizing Antibody Geometric Mean Titers Against Ancestral SARS-CoV-2 (D614G) and Omicron after 50-µg of mRNA-1273.214 and mRNA-1273 Administered as Second Booster Doses by Prior SARS-CoV-2 Infection at Pre-Booster**

| | **All participants** | | **No prior SARS-CoV-2 infection** | | **Prior SARS-CoV-2 infection** | |
|---|---|---|---|---|---|---|
| | **mRNA-1273.214 50 µg Booster Dose** | **mRNA-1273 50 µg Booster Dose** | **mRNA-1273.214 50 µg Booster Dose** | **mRNA-1273 50 µg Booster Dose** | **mRNA-1273.214 50 µg Booster Dose** | **mRNA-1273 50 µg Booster Dose** |
| **Ancestral SARS-CoV-2** (D614G) | **N=428** | **N=367** | **N=334** | **N=260** | **N=94** | **N=98** |
| Pre-booster | | | | | | |
| Observed GMT (95% CI) | 1603.4 (1420.3-1810.0) | 1944.8 (1725.4-2192.1) | 1266.7 (1120.2-1432.5) | 1521.0 (1352.8-1710.2) | 3704.0 (2793.2-4911.7) | 3638.0 (2742.0-4826.6) |
| Day 29 | | | | | | |
| Observed GMT (95% CI) | 6619.0 (5941.7-7373.5) | 6047.5 (5465.9-6691.0) | 5977.3 (5321.9-6713.3) | 5649.3 (5056.8-6311.2) | 9509.7 (7345.9-12,310.9) | 7003.5 (5592.6-8770.4) |
| GMFR (95% CI) | 4.1 (3.8-4.4) | 3.1 (2.9-3.4) | 4.7 (4.4-5.1) | 3.7 (3.4-4.0) | 2.6 (2.2-2.9) | 1.9 (1.6-2.2) |
| **Omicron (B.1.1.529)** | **N=428** | **N=367** | **N=334** | **N=260** | **N=94** | **N=98** |
| Pre-booster | | | | | | |
| Observed GMT (95% CI) | 432.1 (372.5-501.2) | 512.0 (433.4, 604.8) | 298.1 (258.8-343.5) | 332.0 (282.0-390.9) | 1614.6 (1149.7-2267.7) | 1558.4 (1088.9-2230.1) |
| Day 29 | | | | | | |
| Observed GMT (95% CI) | 3070.4 (2685.4-3510.6) | 1932.8 (1681.2-2222.0) | 2372.4 (2070.6-2718.2) | 1473.5 (1270.8-1708.4) | 7676.2 (5618.2-10488.1) | 3885.6 (2877.8-5246.4) |
| GMFR (95% CI) | 7.1 (6.5-7.8) | 3.8 (3.4-4.2) | 8.0 (7.2-8.8) | 4.4 (4.0-5.0) | 4.8 (4.0-5.7) | 2.5 (2.1-3.0) |

SARS-CoV-2 infection baseline info missing for 9 participants in mRNA-1273 50 µg group. CI = Confidence interval, GMT=geometric mean titer LLOQ, lower limit of quantification ULOQ, upper limit of quantification. Antibody values assessed by pseudovirus neutralizing antibody assay reported as below the LLOQ (18.5 for ancestral SARS-CoV-2 [D614G] and 19.9 for omicron B.1.1.529) are replaced by 0.5 × LLOQ. Values greater than ULOQ (45,118) for ancestral SARS-CoV-2 [D614G] and 15,502.7 for omicron B.1.1.529) are replaced by the ULOQ if actual values are not available. 95% CI is calculated based on the t-distribution of the log-transformed values or the difference in the log-transformed values for GM value and GM fold-rise, respectively, then back transformed to the original scale.

**Table S8. Observed Neutralizing Antibody Geometric Mean Titers Against Omicron BA.4 and BA.5 Subvariants after 50-µg of mRNA-1273.214 Administered as Second Booster Doses by Prior SARS-CoV-2 Infection at Pre-Booster**

| | All participants<br>N=428 | No prior SARS-CoV-2 infection<br>N=334 | Prior SARS-CoV-2 infection<br>N=94 |
|---|---|---|---|
| Pre-booster n† | 428 | 334 | 94 |
| Observed GMT (95% CI)§ | 172.7<br>(147.4- 202.3) | 115.6<br>(98.5- 135.6) | 719.5<br>(531.6- 973.9) |
| Day 29, n† | 427 | 333 | 94 |
| Observed GMT (95% CI)§ | 940.6<br>(826.3-1070.6) | 727.4<br>(632.8-836.1) | 2337.4<br>(1825.5-2992.9) |
| GMFR (95% CI)§ | 5.4 (5.0-5.9) | 6.3 (5.7-6.9) | 3.2 (2.8-3.8) |

CI = confidence interval, GM = geometric mean, GMFR = geometric mean fold rise (post-baseline/baseline titers).
n = Number of subjects with non-missing data at baseline and the corresponding post-baseline timepoint.
Antibody values assessed by a research-grade pseudovirus neutralizing antibody ID50 assay for omicron (BA.4/BA.5) reported as below the lower limit of detection ([LOD] 10) are replaced by 0.5 x LOD.
†Number of participants with non-missing data at the timepoint (baseline or post-baseline).
§95% CI is calculated based on the t-distribution of the log-transformed values or the difference in the log-transformed values for GM value and GMFR, respectively, then back transformed to the original scale for presentation.

**Table S9. Binding Antibody Titers and Geometric Mean Ratios Against Ancestral SARS-CoV-2 and Variants after 50-µg of mRNA-1273.214 and mRNA-1273 Administered as Second Booster Doses in Participants with No Evidence of Prior SARS-CoV-2 Infection at Pre-Booster**

| | Ancestral SARS-CoV-2 (D614G) | | Omicron | | Beta | |
|---|---|---|---|---|---|---|
| | mRNA-1273.214 50 µg N=334 | mRNA-1273 50 µg N=260 | mRNA-1273.214 50 µg N=334 | mRNA-1273 50 µg N=260 | mRNA-1273.214 50 µg N=334 | mRNA-1273 50 µg N=260 |
| Pre-booster, n† | 332 | 259 | 331 | 256 | 332 | 259 |
| Observed GM levels§ | 253,732 (230,053-279,847) | 280,187 (255,546-307,204) | 50,656 (45,583-56,293) | 55,962 (50,769-61,686) | 113,986 (103,401-125,656) | 125,812 (114,408-138,352) |
| Day 29, n† | 313 | 251 | 292 | 239 | 306 | 252 |
| Observed GM Level (95% CI)§ | 850,554 (780,987-926,317) | 783,449 (714,773-858,725) | 201,643 (183,823-221,191) | 173,721 (157,525-191,582) | 393467 (362,016- 427,651) | 363114 (331,815-397,366) |
| GMFR (95% CI)§ | 3.4 (3.2-3.6) | 2.8 (2.7-3.0) | 4.0 (3.7-4.2) | 3.1 (2.9-3.4) | 3.5 (3.3- 3.7) | 3.0 (2.8-3.2) |
| Estimated GM Level (95% CI)¶ | 884,978 (839,674-932,727) | 761,984 (718,961-807,582) | 209,859 (198,170-222,237) | 168,687 (158,454-179,580) | 408,455 (387,526-430,513) | 354,483 (334,714-375,419) |
| GMR (95% CI)¶ | 1.16 (1.07-1.26) | | 1.24 (1.14-1.35) | | 1.15 (1.07-1.25) | |
| | **Delta** | | **Gamma** | | **Alpha** | |
| | **mRNA-1273.214 50 µg N=334** | **mRNA-1273 50 µg N=260** | **mRNA-1273.214 50 µg N=334** | **mRNA-1273 50 µg N=260** | **mRNA-1273.214 50 µg N=334** | **mRNA-1273 50 µg N=260** |
| Pre-booster, n† | 332 | 259 | 332 | 259 | 332 | 259 |
| Observed GM levels§ | 160,021 (145,267- 176,274) | 176,051 (160,245-193,415) | 121,087 (109,783-133,556) | 136,347 (124,199-149,684) | 183,545 (166,029-202,909) | 207,233 (188,611-227,693) |
| Day 29, n† | 310 | 252 | 307 | 252 | 308 | 250 |
| Observed GM Level (95% CI)§ | 527,855 (486,349-572,904) | 507,471 (465,240- 553,536) | 424,440 (389,001-463,108) | 386785 (353150- 423624) | 638,782 (585,808-696,546) | 584,312 (532,975-640,594 |
| GMFR (95% CI)§ | 3.3 (3.1-3.5) | 3.0 (2.8-3.1) | 3.5(3.3-3.8) | 2.9 (2.7-3.1) | 3.5 (3.3- 3.7) | 2.9 (2.7-3.1) |
| Estimated GM Level (95% CI)¶ | 546,567 (520,178-574,295) | 494,689 (468,536-522,302) | 443,655 (420,473, 468,114) | 374,273 (352,956-396,878) | 668,710 (634,902-704,319) | 562,715 (531,544-595,715) |
| GMR (95% CI)¶ | 1.11 (1.03-1.19) | | 1.19 (1.10-1.28) | | 1.19 (1.10-1.28) | |

CI = confidence interval, GM = geometric mean, GMR = geometric mean ratio (mRNA-1273.214 relative to mRNA-1273), LLOQ, lower limit of quantification; Meso Scale Discovery =MSD; ULOQ, upper limit of quantification. Binding antibody values assessed by MSD assay reported as below the LLOQ (69 for ancestral SARS-CoV-2; 102 for omicron, 111 for beta; 150 for delta; 143 for gamma; and 52 for alpha) were replaced by 0.5 × LLOQ. Values greater than ULOQ (14,400,000 for ancestral SARS-CoV-2; 1,180,000 for omicron; 5,000,000 for beta; 8,000,000 for delta; 5,800,000 for gamma; 8,800,000 for alpha) were replaced by the ULOQ if actual values are not available.
†Number of participants with non-missing data at the timepoint (baseline or post-baseline).
§95% CI based on the t-distribution of log-transformed values or difference in the log-transformed values for GMT value and GMT fold-rise, respectively, then back transformed to the original scale.
¶Log-transformed antibody levels are analyzed using an ANCOVA model with the treatment variable as fixed effect, adjusting for age group (<65, ≥65 years) and pre-booster titers. The resulting LS means, difference of LS means, and 95% CI are back transformed to the original scale for presentation.

**Table S10. Binding Antibody Levels and Geometric Mean Ratios Against Ancestral SARS-CoV-2 and Variants after 50-µg of mRNA-1273.214 and mRNA-1273 Administered as Second Booster Doses, Regardless of Evidence of Prior SARS-CoV-2 Infection**

| | Ancestral SARS-CoV-2 | | Omicron | | Beta | |
|---|---|---|---|---|---|---|
| | **mRNA-1273.214 50 µg N=428** | **mRNA-1273 50 µg N=367** | **mRNA-1273.214 50 µg N=428** | **mRNA-1273 50 µg N=367** | **mRNA-1273.214 50 µg N=428** | **mRNA-1273 50 µg N=367** |
| Pre-booster, n† | 423 | 365 | 420 | 360 | 423 | 365 |
| Observed GM levels§ | 282,635 (257,514-310,207) | 299,028 (274,090-326,237) | 54,956 (49,719-60,745) | 58,640 (53,339-64,469) | 127,083 (115,948-139,288) | 135,259 (123,818-147,756) |
| Day 29, n† | 405 | 353 | 384 | 334 | 398 | 350 |
| Observed GM Level (95% CI)§ | 856,394 (791,569-926,528) | 753,438 (694,045-817,914) | 201,367 (185,010-219,170) | 163,486 (149,305-179,013) | 403,166 (373,553-435,126) | 352,739 (325,581-382,162) |
| GMFR (95% CI)§ | 3.0 (2.8-3.2) | 2.6 (2.4-2.7) | 3.6 (3.4-3.8) | 2.8 (2.7-3.0) | 3.1 (3.0-3.3) | 2.7 (2.6-2.9) |
| Estimated GM Level (95% CI)¶ | 797,188 (758,939-837,365) | 700,606 (665934-737,084) | 187,826 (178,054-198,134) | 152,403 (144,175-161,100) | 382,058 (363,634-401,415) | 334,638 (317,957-352,195) |
| GMR (95% CI)¶ | 1.14 (1.07-1.21) | | 1.23 (1.15-1.32) | | 1.14 (1.07-1.22) | |
| | **Delta** | | **Gamma** | | **Alpha** | |
| | **mRNA-1273.214 50 µg N=428** | **mRNA-1273 50 µg N=367** | **mRNA-1273.214 50 µg N=428** | **mRNA-1273 50 µg N=367** | **mRNA-1273.214 50 µg N=428** | **mRNA-1273 50 µg N=367** |
| Pre-booster, n† | 423 | 365 | 423 | 365 | 423 | 364 |
| Observed GM levels§ | 181,213 (165,495-198,424) | 193,939 (177,757-211,594) | 138,574 (126,336-151,998) | 150,030 (137,680-163,486) | 207,587 (188,619-228,462) | 223,595 (204,443-244,541) |
| Day 29, n† | 402 | 351 | 399 | 350 | 400 | 350 |
| Observed GM Level (95% CI)§ | 546,450 (507,230-588,702) | 502,005 (465,032-541,917) | 441,810 (408,370-477,989) | 384,401 (354,977-416,264) | 652,098 (602,056- 706,299) | 568,576 (523,291-617,779) |
| GMFR (95% CI)§ | 3.0 (2.8-3.2) | 2.7 (2.5-2.8) | 3.1 (3.0-3.3) | 2.7 (2.5-2.8) | 3.1 (2.9-3.3) | 2.6 (2.5-2.7) |
| Estimated GM Level (95% CI)¶‖ | 516,916 (493,390-541,564) | 472,237 (450,021-495,549 | 419,813 (399,165-441,528) | 361,839 (343,419- 381,246)) | 610,201 (581,170, 640,682) | 523,953 (498,178-551,061) |
| GMR (95% CI)¶‖ | 1.10 (1.03-1.16) | | 1.16 (1.09-1.24) | | 1.17 (1.09-1.24) | |

ANCOVA=analysis of covariance, CI = Confidence interval, GM=geometric mean, GMR = Geometric mean ratio, mRNA-1273.214 vs mRNA-1273. LLOQ, lower limit of quantification; LS, least squares; ULOQ, upper limit of quantification. Binding antibody values assessed by MSD assay reported as below the LLOQ (69 for ancestral SARS-CoV-2; 102 for omicron, 111 for beta; 150 for delta; 143 for gamma; and 52 for alpha) were replaced by 0.5 × LLOQ. Values greater than ULOQ (14,400,000 for ancestral SARS-CoV-2; 1,180,000 for omicron; 5,000,000 for beta; 8,000,000 for delta; 5,800,000 for gamma; 8,800,000 for alpha) were replaced by the ULOQ if actual values are not available.
†Number of participants with non-missing data at the timepoint (baseline or post-baseline).
§95% CI based on the t-distribution of log-transformed values or difference in the log-transformed values for GMT value and GMT fold-rise, respectively, then back transformed to the original scale.
¶Log-transformed antibody levels are analyzed using an ANCOVA model with the treatment variable as fixed effect, adjusting for age group (<65, ≥65 years), pre-booster titers and prior SARS-CoV-2 infection. The resulting LS means, difference of LS means, and 95% CI are back transformed to the original scale.

**Table S11. Observed Binding Antibody Geometric Mean Levels Against Ancestral SARS-CoV-2 and Omicron After Second 50-µg Boosters of mRNA-1273.214 and mRNA-1273 by Prior SARS-CoV-2 Infection at Pre-Booster**

| | All participants | | No prior SARS-CoV-2 infection | | Prior SARS-CoV-2 infection | |
|---|---|---|---|---|---|---|
| | mRNA-1273.214 50 µg Booster Dose | mRNA-1273 50 µg Booster Dose | mRNA-1273.214 50 µg Booster Dose | mRNA-1273 50 µg Booster Dose | mRNA-1273.214 50 µg Booster Dose | mRNA-1273 50 µg Booster Dose |
| **Ancestral SARS-CoV-2** | **N=428** | **N=367** | **N=334** | **N=260** | **N=94** | **N=98** |
| Pre-booster | | | | | | |
| n | 423 | 365 | 332 | 259 | 91 | 97 |
| Observed GM level (95% CI) | 282,635 (257,514-310207) | 299,028 (274,090-326,237) | 253,732 (230,053-279,847) | 280,187 (255,546-307,204) | 418,947 (332,856-527,305) | 349,738 (283,191-431,922) |
| Day 29 | | | | | | |
| n | 405 | 353 | 313 | 251 | 92 | 95 |
| Observed GM level (95% CI) | 856,394 (791,569-926,528) | 753,438 (694,045-817,914) | 850,554 (780,987-926,317) | 783,449 (714,773-858,725) | 876,567 (723,100-1,062,604) | 682,474 (567,428-820,846) |
| GMFR (95% CI) | 3.0 (2.8-3.2) | 2.6 (2.4-2.7) | 3.4 (3.2-3.6) | 2.8 (2.7-3.0) | 2.0 (1.8-2.2) | 1.9 (1.8-2.1) |
| **Omicron (B.1.1.529)** | **N=428** | **N=367** | **N=334** | **N=260** | **N=94** | **N=98** |
| Pre-booster | | | | | | |
| n | 420 | 360 | 331 | 256 | 89 | 95 |
| Observed GM level (95% CI) | 54,956 (49,719-60,745) | 58,640 (53,339-64,469) | 50,656 (45,583-56,293) | 55,962 (50,769-61,686) | 74,410 (57,476-96,333) | 65,018 (51,139-82,664) |
| Day 29 | | | | | | |
| n | 384 | 334 | 292 | 239 | 92 | 88 |
| Observed GM level (95% CI) | 201,367 (185,010-219,170) | 163,486 (149,305-179,013) | 201,643 (183,823-221,191) | 173,721 (157,525-191,582) | 200,491 (164,075-244,991) | 139,392 (112,194-173,182) |
| GMFR (95% CI) | 3.6 (3.4-3.8) | 2.8 (2.7-3.0) | 4.0 (3.7-4.2) | 3.1 (2.9-3.4) | 2.5 (2.3-2.8) | 2.2 (2.0-2.4) |

SARS-CoV-2 infection baseline info missing for 9 participants in mRNA-1273 50 µg group. CI = Confidence interval, GM=geometric mean, GSLM=geometric least squares mean, GMR=geometric mean ratio, LLOQ, lower limit of quantification. Meso Scale Discovery=MSD; ULOQ, upper limit of quantification. Binding IgG antibody values assessed by MSD assay reported as below the LLOQ (69 for ancestral SARS-CoV-2; 102 for omicron B.1.1.529) are replaced by 0.5 × LLOQ. Values greater than ULOQ (14,400,000 for ancestral SARS-CoV-2; 1,180,000 for omicron) are replaced by the ULOQ if actual values are not available. 95% CI was calculated based on the t-distribution of the log-transformed values or the difference in the log-transformed values for GM value, and GM fold-rise, (and GLSM and GMR), respectively, and then back transformed to the original scale for presentation.
