## Supplementary material for "A Bivalent Omicron-containing Booster Vaccine Against Covid-19": CONSORT Checklist

### CONSORT Statement 2006 - Checklist for Non-inferiority and Equivalence Trials

**Items to include when reporting a non-inferiority or equivalence randomized trial**

**Please note: This is an open-label phase 2/3 study that evaluates safety and non-inferiority of study vaccination in 2 cohorts of participants. The checklist has been correspondingly completed to indicate compliance with guidelines where applicable. NA=not applicable.**

| ***PAPER SECTION*** **And topic** | **Item** | **Descriptor** | **Reported on Page #** |
|---|---|---|---|
| *TITLE & ABSTRACT* | 1 | How participants were allocated to interventions (*e.g.*, "random allocation", "randomized", or "randomly assigned"), *specifying that the trial is a non-inferiority or equivalence trial.* | NA: not a randomized trial; participants were allocated to study vaccine as described (pg. 5, & Fig. 1, pg.19) |
| *INTRODUCTION* Background | 2 | Scientific background and explanation of rationale, *including the rationale for using a non-inferiority or equivalence design.* | 4; Supplement pg.4-6 |
| *METHODS* Participants | 3 | Eligibility criteria for participants *(detailing whether participants in the non-inferiority or equivalence trial are similar to those in any trial(s) that established efficacy of the reference treatment)* and the settings and locations where the data were collected. | 5; Supplement, pg. 4-6 |
| Interventions | 4 | Precise details of the interventions intended for each group *detailing whether the reference treatment in the non-inferiority or equivalence trial is identical (or very similar) to that in any trial(s) that established efficacy,* and how and when they were actually administered. | 5; Supplement pg. 4 |
| Objectives | 5 | Specific objectives and hypotheses, *including the hypothesis concerning non-inferiority or equivalence.* | 7-9; Supplement, pg. 6-9 and 16-17 |
| Outcomes | 6 | Clearly defined primary and secondary outcome measures *detailing whether the outcomes in the non-inferiority or equivalence trial are identical (or very similar) to those in any trial(s) that established efficacy of the reference treatment* and, when applicable, any methods used to enhance the quality of measurements (*e.g.*, multiple observations, training of assessors). | 7-9; Supplement, pg. 6-9 |
| Sample size | 7 | How sample size was determined *detailing whether it was calculated using a non-inferiority or equivalence criterion and specifying the margin of equivalence with the rationale for its choice*. When applicable, explanation of any interim analyses and stopping rules (*and whether related to a non-inferiority or equivalence hypothesis*). | Supplement, pg. 7 |
| Randomization -- Sequence generation | 8 | Method used to generate the random allocation sequence, including details of any restrictions (*e.g.*, blocking, stratification) | NA: not randomized |
| Randomization -- Allocation concealment | 9 | Method used to implement the random allocation sequence (*e.g.*, numbered containers or central telephone), clarifying whether the sequence was concealed until interventions were assigned. | NA: not randomized |
| Randomization -- Implementation | 10 | Who generated the allocation sequence, who enrolled participants, and who assigned participants to their groups. | NA: not randomized |
| Blinding (masking) | 11 | Whether or not participants, those administering the interventions, and those assessing the outcomes were blinded to group assignment. If done, how the success of blinding was evaluated. | NA: open-label trial |
| Statistical methods | 12 | Statistical methods used to compare groups for primary outcome(s), *specifying whether a one or two-sided confidence interval approach was used.* Methods for additional analyses, such as subgroup analyses and adjusted analyses. | 7-9; Supplement, pg. 6-8 |

| | | | |
|---|---|---|---|
| *RESULTS*<br>Participant flow | 13 | Flow of participants through each stage (a diagram is strongly recommended). Specifically, for each group report the numbers of participants randomly assigned, receiving intended treatment, completing the study protocol, and analyzed for the primary outcome. Describe protocol deviations from study as planned, together with reasons. | 16; Figure 1 |
| Recruitment | 14 | Dates defining the periods of recruitment and follow-up. | 9-10 |
| Baseline data | 15 | Baseline demographic and clinical characteristics of each group. | 9 and Table 1, pg. 20 |
| Numbers analyzed | 16 | Number of participants (denominator) in each group included in each analysis and whether the analysis was *"intention-to-treat" and/or alternative analyses were conducted*. State the results in absolute numbers when feasible (*e.g.*, 10/20, not 50%). | Table 2 pg. 21; Supplement, pg. 12-15 and 22-28 |
| Outcomes and estimation | 17 | For each primary and secondary outcome, a summary of results for each group, and the estimated effect size and its precision (*e.g.*, 95% confidence interval). *For the outcome(s) for which non-inferiority or equivalence is hypothesized, a figure showing confidence intervals and margins of equivalence may be useful*. | 11-12, Table 2; 21; Supplement, p. 12-15 and 19-28 |
| Ancillary analyses | 18 | Address multiplicity by reporting any other analyses performed, including subgroup analyses and adjusted analyses, indicating those pre-specified and those exploratory. | 7-9; Supplement, pg. 6-8 |
| Adverse events | 19 | All important adverse events or side effects in each intervention group. | 10 and Figure 2 pg.17 |
| *DISCUSSION*<br>Interpretation | 20 | Interpretation of the results, taking into account the *non-inferiority or equivalence hypothesis and any other* study hypotheses, sources of potential bias or imprecision and the dangers associated with multiplicity of analyses and outcomes. | 9-12 |
| Generalizability | 21 | Generalizability (external validity) of the trial findings. | 14-15 |
| Overall evidence | 22 | General interpretation of the results in the context of current evidence. | 13-14 |
